# Clinical progression parameters associated with SARS-CoV-2, influenza, and respiratory syncytial virus infections

**DOI:** 10.1101/2025.05.28.25328531

**Authors:** Noah T. Parker, Vennis Hong, Gregg S. Davis, Magdalena Pomichowski, Iris A. Reyes, Fagen Xie, Nicola F. Mueller, Isabel Rodriguez-Barraquer, Sara Y. Tartof, Joseph A. Lewnard

## Abstract

Mathematical and computational models are often used to forecast respiratory infectious disease burden, including to inform healthcare capacity requirements. We aimed to characterize pathways of clinical progression associated with SARS-CoV-2, influenza, and respiratory syncytial virus (RSV) infections using data from patients in an integrated healthcare system, whose encounters were monitored across all levels of acuity spanning virtual, ambulatory, and inpatient care settings. Using parametric survival models, we estimated probabilities of progression and distributions of time to progression from each care setting to all higher-acuity settings on a cascade encompassing the following classes of events or healthcare encounters: symptoms onset; diagnostic testing; telehealth or other virtual care appointment; outpatient physician office visit; urgent care presentation; emergency department presentation; hospital admission; mechanical ventilation; and death. Our analyses included data from 59,668, 22,705, and 1,668 episodes associated with positive SARS-CoV-2, influenza, and RSV tests, respectively, between 1 April 2023 and 31 March 2024. First clinical encounters occurred in inpatient settings for only 4.7%, 3.4%, and 18.7% of SARS-CoV-2, influenza, and RSV episodes, respectively, with median times (interquartile range) of 6.8 (3.6-13.2), 6.6 (3.5-12.1), and 6.4 (3.8-10.6) days from symptoms onset to admission. Overall, 7.9% of SARS-CoV-2 episodes, 5.8% of influenza episodes, and 33.8% of RSV episodes resulted in inpatient admission, ventilation, or death. Between 40.4-62.1%, 71.6-87.3%, and 47.9-58.7% of SARS-CoV-2, influenza, and RSV infections, respectively, had encounters in lower-acuity virtual care, outpatient, or urgent care settings. For all three viruses, the proportions of cases receiving care at each level of acuity increased with older age and greater numbers of comorbid conditions. Median durations of hospital stay were 4.2 (2.6-7.3), 4.0 (2.3-6.8), and 4.3 (2.5-7.4) days for SARS-CoV-2, influenza, and RSV episodes resulting in admission. These estimates provide a basis for modeling real-world clinical care requirements and the progression of respiratory viral infections.

**AUTHOR SUMMARY:** Models of respiratory infections such as SARS-CoV-2, influenza, and RSV are used to forecast disease burden and plan the allocation of healthcare resources. Early healthcare system encounters—for instance, for receipt of care in virtual or outpatient settings—may provide a basis for anticipating higher-acuity care requirements as patients progress to more severe disease. However, limited data are available addressing patterns of healthcare utilization among patients with these infections. Using electronic healthcare records from an integrated healthcare system, we estimated probabilities and rates of progression from lower-acuity states, such as virtual or outpatient visits, to increasingly higher-acuity states including inpatient admission, ventilation, and death. We quantified associations of demographic and clinical risk factors with progression probabilities for each infection. We provide a databank containing fitted distributions for progression to inform infectious disease modeling.

## INTRODUCTION

Acute respiratory illnesses (ARIs) caused by SARS-CoV-2, influenza, and respiratory syncytial virus (RSV) are important contributors to morbidity and mortality in the United States and globally (1–3). Anticipating healthcare utilization associated with ARIs is an objective of both public health agencies and healthcare delivery organizations. Mathematical and computational models used to forecast ARI burden are often trained using data from either syndromic surveillance or reported cases, hospital admissions, and deaths associated with each infection (4). To inform capacity planning— including decisions around the allocation of personnel, space, medications, laboratory infrastructure, and other resources—such models require realistic parameters concerning the likelihood and time course of cases’ healthcare utilization across settings of varying acuity.

Despite this need, few real-world data sources address clinical care trajectories associated with ARIs due to SARS-CoV-2, influenza, and RSV. While parameters such as the proportion of SARS-CoV-2 infections resulting in hospital admission or death (5,6) and durations of hospital stay (7,8) were estimated in numerous settings during the early phases of the COVID-19 pandemic, fewer studies have addressed utilization patterns in lower-acuity healthcare settings such as ambulatory clinics and emergency departments, where the greatest numbers of all medically-attended cases receive care. Moreover, updated epidemiological parameter estimates for recent SARS-CoV-2 variants and in populations with widespread immunity are not widely available. These challenges are equally pronounced in efforts to model seasonal influenza and RSV. Although some studies have aimed to characterize reporting pyramids addressing symptomatic or medically attended, hospitalized, and fatal influenza cases for contexts of seasonal (9) and pandemic influenza (10,11), these studies have drawn on data from disparate sources and settings, and do not address time-to-event parameters that are likewise critical to forecasting.

We aimed to characterize pathways of clinical care progression during ARIs associated with SARS-CoV-2, influenza, and RSV. We analyzed data from patients enrolled in capitated, managed care plans within an integrated healthcare system in southern California. This rich data source allowed us to monitor patient encounters across all levels of acuity spanning virtual, ambulatory, and inpatient care settings. We quantified how patients progress through the different settings of care using parametric survival models among all ascertained infections. The outputs of this analysis provide a basis for modeling clinical burden and healthcare system impacts of SARS-CoV-2, influenza, and RSV infections.

## METHODS

### Overview of the modeling approach

We defined a cascade of clinical progression wherein we aimed to estimate (a) the probability that an individual observed at any state along this cascade would progress to a higher-acuity state, and (b) the distributions of times to progression from lower-acuity to higher-acuity states (**Figure 1**). Current states were characterized as the highest-acuity settings where individuals had received care at a given point during their infection. We defined the states in order of increasing acuity as: any infection or symptomatic infection without associated healthcare utilization (besides testing); telehealth or other virtual care appointment; outpatient physician office visit; urgent care presentation; emergency department presentation; hospital admission; mechanical ventilation; and death. Our analyses account for the fact that individuals may progress from lower-to higher-acuity states without being intercepted via healthcare encounters at intermediate levels between these origin and destination states.

**Figure 1:**
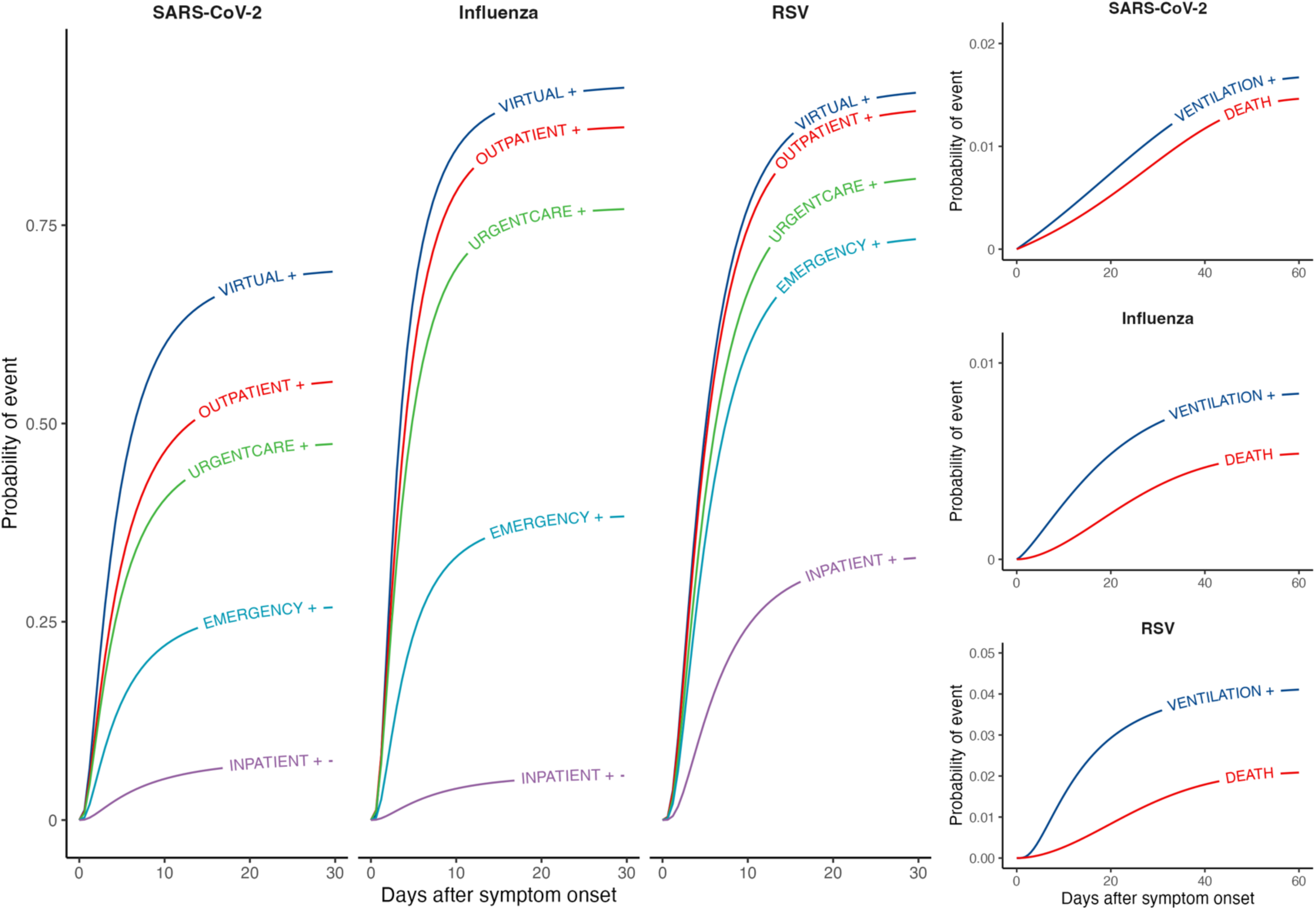
Clinical care cascade for acute respiratory illnesses associated with SARS-CoV-2, influenza, and RSV infections. We illustrate cumulative distribution functions from best-fitting models (defined by Akaike information criterion values) for times from symptom onset to progression to (or beyond) each acuity threshold. Right-hand panels illustrate cumulative distribution functions for rare outcomes (mechanical ventilation and death).

We fit parametric survival models to jointly estimate the probabilities of progression, and the distributions of time to progression, from each originating state to all higher-acuity states on the cascade. We modeled three classes of transitions aiming to inform distinct forecasting applications. First, we modeled an individual’s most proximal progression event from each originating state on the cascade, aiming to characterize typical pathways of care utilization. Secondly, we modeled an individual’s total probability of progression to each state or higher-acuity states from all originating states on the cascade, aiming to inform projections of total demand at each level of acuity within a patient cohort observed at any point in time. Lastly, we aimed to estimate durations of hospital stay following inpatient admission. We explored the association of progression risk and progression rates with individual epidemiologic and demographic characteristics by including covariates in survival models.

We share parameters of all fitted models for re-use within the supplementary materials and via an online repository (https://github.com/ntparker3/Resp_params).

### Setting

We used data from healthcare encounters among members of Kaiser Permanente Southern California (KPSC), an integrated healthcare system providing care across virtual, outpatient, and inpatient settings to roughly 4.7 million individuals throughout southern California during the study period. Members of KPSC enroll through a combination of employer-sponsored, pre-paid, and government-subsidized insurance plans, and broadly reflect the socioeconomic and racial and ethnic composition of the area’s population (12,13). Electronic health records (EHRs) capture clinical notes, diagnoses, laboratory results, and prescriptions for care received at KPSC facilities, while insurance claims capture out-of-network care, enabling near-complete ascertainment of healthcare delivery for members.

### Study population and episode definition

We conducted event-level analyses of ARIs associated with SARS-CoV-2, influenza, and RSV among individuals of any age who tested positive for each pathogen by molecular or antigen-based assays in any clinical setting between 1 April, 2023 and 31 March, 2024. We selected this study period to ensure results were not impacted by disruptions in routine care delivery associated with earlier emergency phases of the COVID-19 pandemic. While co-infections were exceptionally rare, we allowed overlapping episode periods to contribute to observations for each unique virus identified. We limited the study population to individuals with ≥1 year of continuous enrollment before their index test (allowing for enrollment gaps up to 45 days in length) to ensure accurate characterization of individuals’ baseline health status from prior-year utilization.

We defined index tests for each ARI episode as the date of specimen collection associated with the first positive test for each pathogen during the study period. Symptoms data (presence of and onset dates for fever, cough, headache, fatigue, dyspnea, chills, sore throat, myalgia, anosmia, diarrhea, vomiting or nausea, and abdominal pain within 14 days before testing) were solicited at the point of testing for all individuals who received SARS-CoV-2 tests during the study period.

Symptoms were recorded most ARI episodes as nearly all individuals who received influenza or RSV tests within KPSC were previously or concurrently tested for SARS-CoV-2. We supplemented structured data from symptoms questionnaires with searches of free-text EHR fields via a previously described natural language processing (NLP) algorithm (14) to characterize the presence and onset times of symptoms using all available information. We defined the date of symptoms onset as the earliest recorded date within 14 days before to 30 days after testing for each episode.

We characterized ARI episodes using data from all healthcare encounters occurring between the date of symptoms onset (or up to seven days before the date of testing, if symptoms were not recorded at the time of testing) and 30 days after the testing date. We defined dates of progression to each state as the first date at which individuals received care in the associated clinical setting with at least one ARI-related diagnosis code recorded (**Table S1**). We excluded care utilization not associated with such codes to ensure that healthcare encounters unrelated to an ongoing ARI episode were not interpreted as indicators of disease progression.

### Statistical analysis

We fit parametric survival models using the *flexsurv* package (15) in R (version 4.4.2; R Foundation for Statistical Computing, Vienna, Austria). We estimated parameters corresponding to assumptions that times-to-event followed Exponential, Gamma, Generalized Gamma, Log Normal, Weibull, and Gompertz distributions for all transitions. We describe results for models with the lowest Akaike information criterion (AIC) values for each transition in this manuscript, and present parameter estimates for all distributions evaluated within the accompanying code base (https://github.com/ntparker3/Resp_params).

For models of the most proximal transition from each state, we followed for progression events within 20 days after dates of entry into each originating state. We considered observations to be interval-censored if no progression event occurred within 20 days. As a sensitivity analysis, we also present estimates for models including follow-up through 60 days for these transitions. We used mixture models to estimate probabilities of and times to the most proximal progression event from each originating state. These models defined distinct rates for progression between each originating state and all higher-acuity states, and handled progression to each state as a competing risk. This framework corresponded to the interpretation that progression to a higher-acuity state of illness could precede progression to intermediate states along the same cascade.

For models of individuals’ total risk of progression to each acuity state (or higher-acuity states), we followed for progression events within 60 days after dates of entry into each originating state. When individuals experienced care corresponding to multiple states on the same day (e.g., an emergency presentation leading to hospital admission), we defined the highest-acuity state observed as the outcome. In contrast to analyses for individuals’ most proximal transition, models for individuals’ total risk of progression to each state—cumulatively across all intermediate pathways of care—did not require a competing-risks framework. For these analyses, we instead recorded progression as occurring when individuals experienced the outcome of interest or one signifying receipt of higher-acuity care.

To estimate hospital lengths of stay, we fit parametric survival models defining admission dates as originating events and dates of discharge or in-hospital mortality as the outcome; we modeled consecutive admissions with same-day discharges and readmissions as continuous hospitalization events. We also used mixture models defining competing risks for death and discharge to separately estimate durations of hospitalization according to individuals’ clinical outcome.

We repeated the analyses described above to estimate subgroup-specific risks or rates of progression based on age, sex, race/ethnicity, vaccination status, Charlson comorbidity index, and community-level socioeconomic status, as measured by census tract-level neighborhood deprivation index values (16). We categorized continuous covariates according to the distributions presented in **Table 1**. We fit parametric survival models allowing variation across covariate strata in both the probability of progression and the location parameter for times-to-event for each modeled distribution. As for our primary analyses, we describe results for models yielding the lowest AIC value for each transition in this manuscript, and present parameter estimates for all distributions in the accompanying repository.

**Table 1:** Individual characteristics, by infecting virus.

| Characteristic |  | Cases, by infecting virus, *n* (%) |  |  |
| --- | --- | --- | --- | --- |
|  |  | SARS-CoV-2<br>*N*=59,668 | Influenza<br>*N*=22,705 | RSV<br>*N*=1,668 |
| Age, years | 0-17 | 4,489 (7.5) | 7,215 (31.8) | 920 (55.2) |
|  | 18-49 | 23,033 (38.6) | 8,261 (36.4) | 71 (4.3) |
|  | 50-59 | 9,307 (15.6) | 2,619 (11.5) | 87 (5.2) |
|  | 60-69 | 8,929 (15.0) | 2,191 (9.6) | 146 (8.8) |
|  | 70-79 | 8,088 (13.6) | 1,612 (7.1) | 176 (10.6) |
|  | 80-89 | 4,639 (7.8) | 662 (2.9) | 193 (11.6) |
|  | ≥90 | 1,183 (2.0) | 145 (0.6) | 75 (4.5) |
| Sex | Male | 24,193 (40.5) | 10,236 (45.1) | 775 (46.5) |
|  | Female | 35,475 (59.5) | 12,469 (54.9) | 893 (53.5) |
| Race/ethnicity | White, non-Hispanic | 14,556 (24.4) | 4,988 (22.0) | 482 (28.9) |
|  | Asian, non-Hispanic | 7,033 (11.8) | 2,333 (10.3) | 201 (12.1) |
|  | Black, non-Hispanic | 6,767 (11.3) | 2,155 (9.5) | 166 (10.0) |
|  | Hispanic (any race) | 28,113 (47.1) | 11,894 (52.4) | 764 (45.8) |
|  | Pacific Islander | 484 (0.8) | 165 (0.7) | — |
|  | Native American/Alaska native | 115 (0.2) | 32 (0.1) | 3 (0.2) |
|  | Other | 859 (1.4) | 396 (1.7) | 14 (0.8) |
|  | Multiple | 214 (0.4) | 122 (0.5) | 8 (0.5) |
|  | Unknown | 1,527 (2.6) | 620 (2.7) | 17 (1.0) |
| Insurance source | Commercial | 30,861 (51.7) | 12,704 (56.0) | 688 (41.2) |
|  | Medicaid | 6,201 (10.4) | 3,223 (14.2) | 299 (17.9) |
|  | Medicare | 13,358 (22.4) | 2,419 (10.7) | 400 (24.0) |
|  | Pre-paid plans | 1,444 (2.4) | 695 (3.1) | 33 (2.0) |
|  | Other | 653 (1.1) | 185 (0.8) | 14 (0.8) |
|  | Unknown | 7,151 (12.0) | 3,479 (15.3) | 234 (14.0) |
| Vaccinations | 1-2 COVID-19 vaccine doses | 11,271 (18.9) | 5,300 (23.3) | 156 (9.4) |
|  | 3+ COVID-19 vaccine doses | 41,453 (69.5) | 10,809 (47.6) | 724 (43.4) |
|  | Seasonal influenza vaccine received | 19,103 (32.0) | 10,775 (47.5) | 381 (22.8) |
|  | RSV vaccine received | 443 (0.7) | 210 (0.9) | 27 (1.6) |
| Neighborhood deprivation index | NDI < −1 | 2,565 (4.3) | 1,018 (4.5) | 78 (4.7) |
|  | −1 ≤ NDI < 0 | 19,632 (32.9) | 7,034 (31.0) | 535 (32.1) |
|  | 0 ≤ NDI < 1 | 20,701 (34.7) | 7,871 (34.7) | 576 (34.5) |
|  | NDI > 1 | 11,097 (18.6) | 4,494 (19.8) | 279 (16.7) |
|  | Unknown | 5,673 (9.5) | 2,288 (10.1) | 200 (12.0) |
| Charlson comorbidity index | 0 | 32,336 (54.2) | 14,912 (65.7) | 834 (50.0) |
|  | 1-2 | 16,319 (27.3) | 5,576 (24.6) | 379 (22.7) |
|  | 3-5 | 7,180 (12.0) | 1,460 (6.4) | 253 (15.2) |
|  | 6+ | 3,833 (6.4) | 757 (3.3) | 202 (12.1) |

### Ethics

This study was reviewed and approved by the KPSC institutional review board, which granted a waiver of informed consent for retrospective analysis of EHR data.

## RESULTS

### Descriptive characteristics

Our analyses included data from 348,958 unique KPSC members who received tests for SARS-CoV-2, influenza, or RSV between 1 April, 2023 and 31 March, 2024, among whom we identified 59,668 episodes associated with positive SARS-CoV-2 test results, 22,705 episodes associated with positive influenza test results, and 1,668 episodes associated with positive RSV test results (**Table 1**). Among these episodes, 602 were associated with coinfections (579 SARS-CoV-2 and influenza coinfections, 11 SARS-CoV-2 and RSV coinfections, and 12 influenza and RSV coinfections). The greatest numbers of SARS-CoV-2 and influenza infections occurred among individuals aged 18-49 years (*n*=23,033 [38.6%] and *n*=8,261 [36.4%], respectively). For influenza and RSV, a considerable number of episodes also occurred among children aged ≤17 years (*n*=7,215 [31.8%] and 920 [55.2%], respectively), while 10.7-26.6% of infections with each pathogen occurred among individuals aged ≥70 years (*n*=13,910 with SARS-CoV-2, *n*=2,419 with influenza, and *n*=444 with RSV). Most episodes involving each pathogen occurred among Hispanic individuals of any race or White, non-Hispanic individuals without comorbid conditions. Across all three pathogens, roughly half (41.2-56.0%) of infections occurred among individuals enrolled in commercial insurance plans, and a plurality (10.4-17.9%) occurred among individuals enrolled in Medicare-sponsored plans. Among SARS-CoV-2 infections, 6,944 (11.6%), 11,271 (18.9%), and 41,453 (69.5%) occurred among individuals who had received 0, 1-2, and ≥3 COVID-19 vaccine doses, cumulatively; 10,775 influenza infections (47.5%) occurred among individuals who had received seasonal influenza vaccination, and few RSV infections (*n*=27; 1.6%) occurred among individuals who were previously vaccinated against RSV.

### Care pathways for SARS-CoV-2

For ARIs associated with SARS-CoV-2 infection, the first clinical encounter following symptoms onset most often occurred in urgent care (18.6%) or emergency department (17.9%) settings, followed by virtual care appointments (10.7%), outpatient office visits (7.2%), and inpatient settings (4.7%; **Table 2**; **Table S3**; **Table S4**). Median time from symptoms onset to testing was 3.2 days (**Figure 2**). Among individuals who received virtual care, 21.1% subsequently received care in higher-acuity clinical settings in the following 20 days, with 7.2%, 5.0%, and 7.1%, presenting to outpatient office visits, urgent care facilities, and emergency departments as their next clinical encounter, respectively; 1.9% were admitted to hospital at their next clinical encounter (**Table 2**). For individuals who were admitted at their next clinical encounter after a virtual care appointment, median time to admission was 2.0 days (interquartile range [IQR]: 0.6-5.2). Among individuals who received care at outpatient and urgent care facilities, 4.0% and 1.7%, respectively, were admitted to the hospital at their next clinical encounter after a median of 0.9 and 0.4 days, respectively.

**Table 2:**
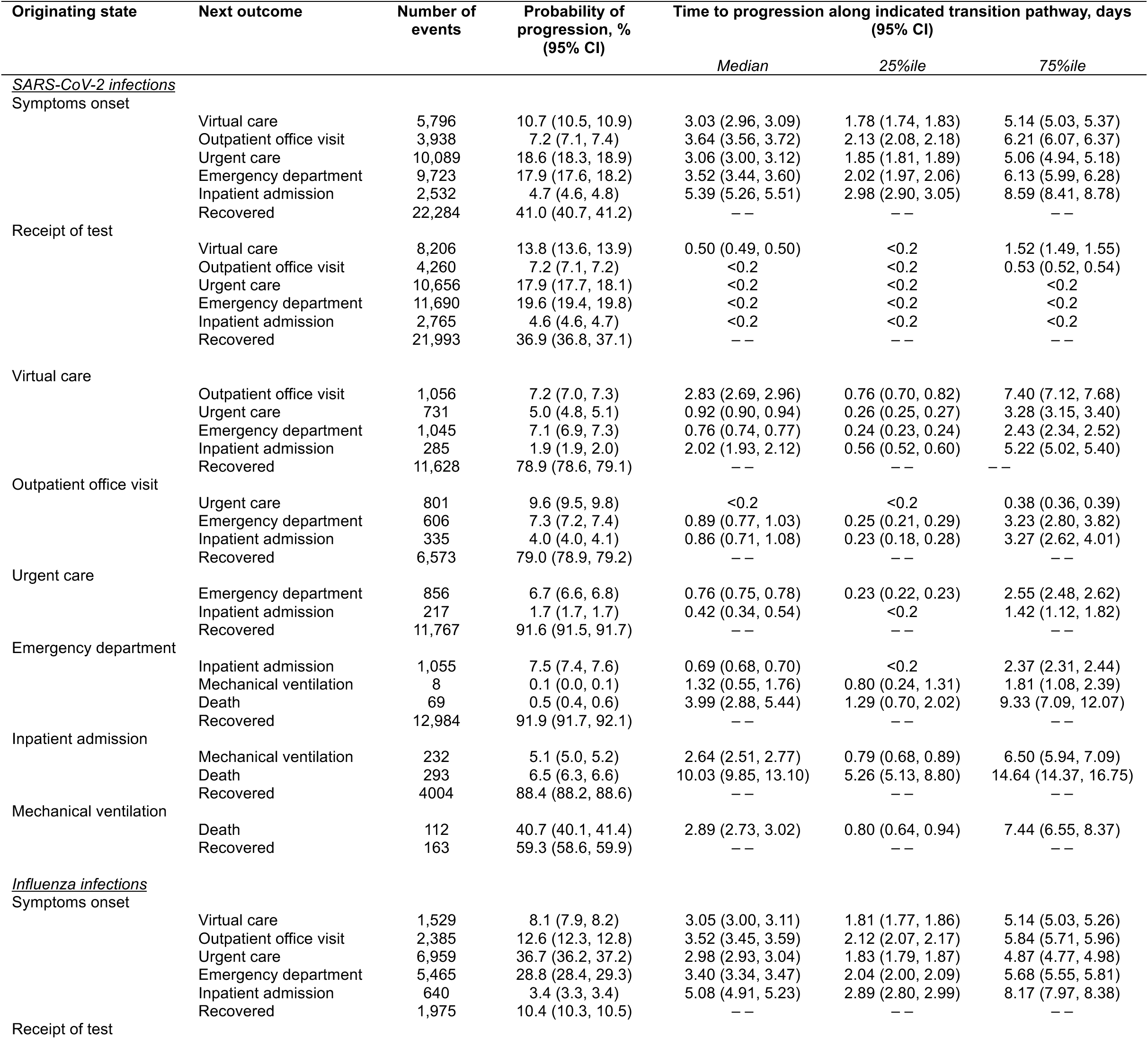
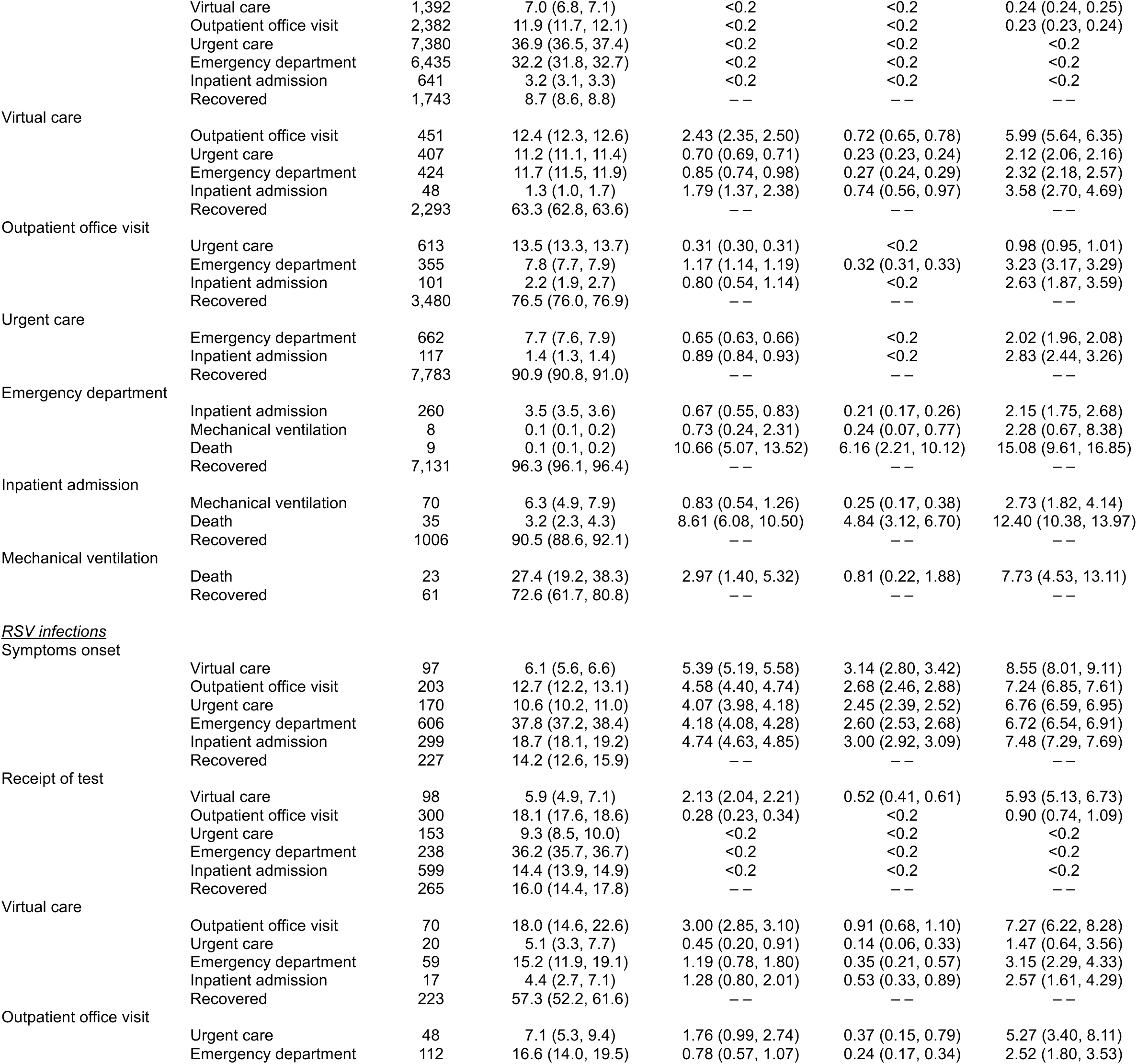
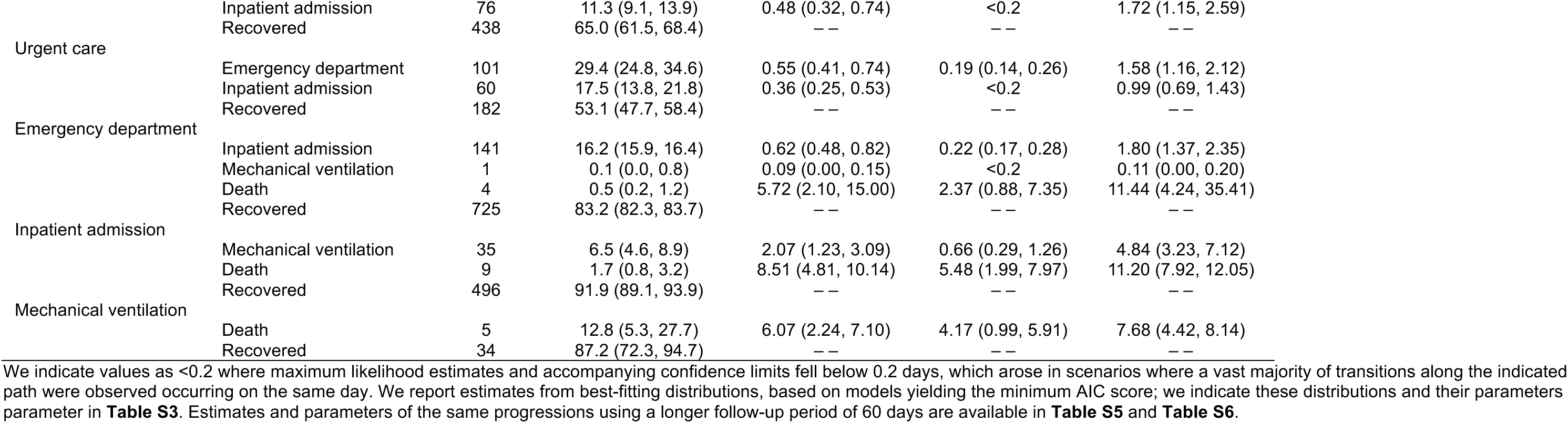
Care utilization pathways associated with each infecting virus using a follow-up period of 20 days.

**Figure 2:**
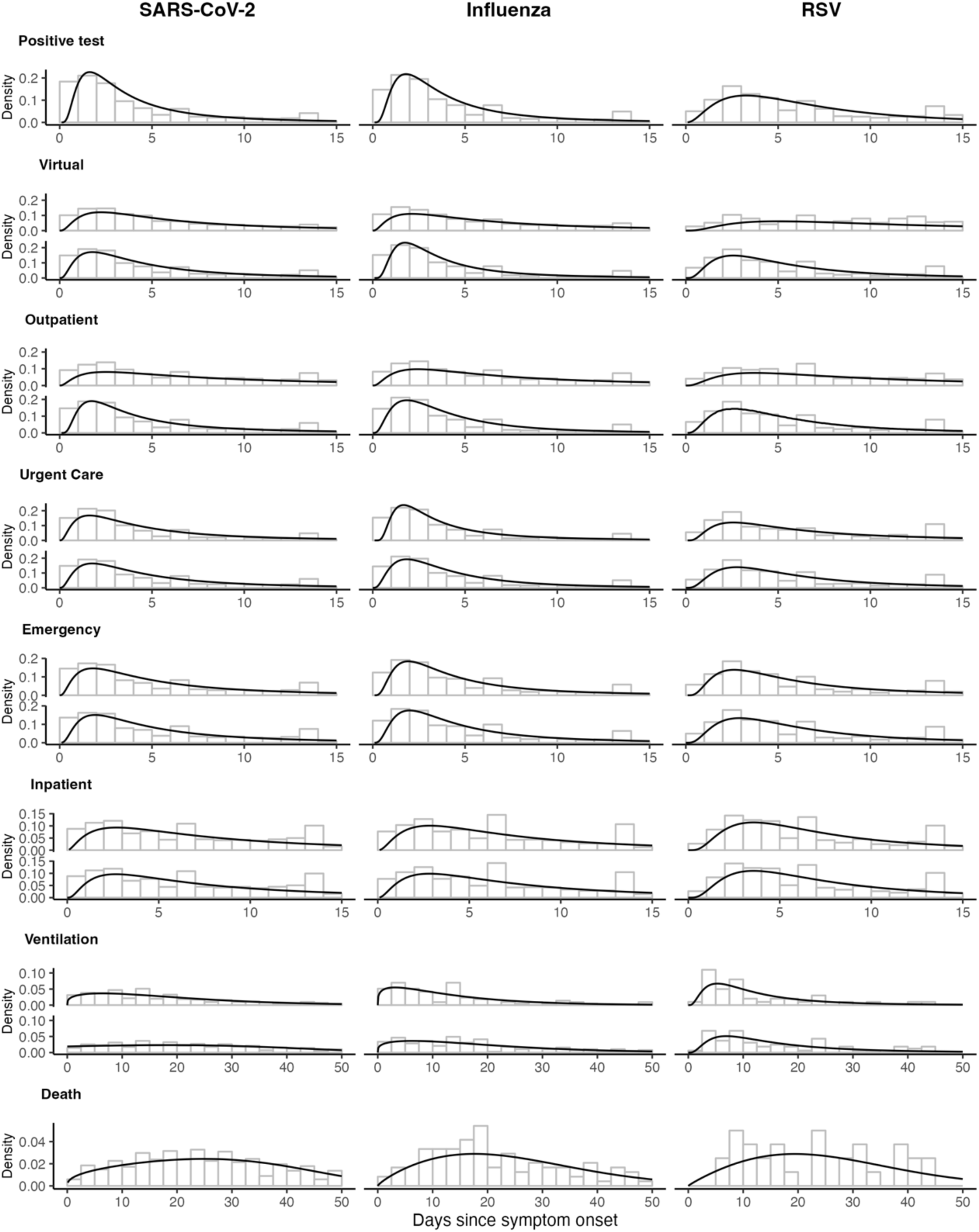
Time-to-event distributions of reaching acuity thresholds from symptom onset for illnesses associated with SARS-CoV-2, influenza, and RSV. For each outcome, top panels illustrate the distribution of times from symptom onset to receipt of care at the indicated state, while bottom panels illustrate the distribution of times from symptom onset to receiving care at or above the indicated acuity threshold. Panels for *positive test* and *death* show the time from symptom onset to reaching the exact state only. Black lines represent the density of the best-fitting distribution, selected by AIC.

Median duration of inpatient stay for SARS-CoV-2 infections was 4.2 days (IQR: 2.6-7.3); median time to discharge was 4.1 days (IQR: 2.6-6.9) for patients who were discharged alive (**Figure 3**; **Table S7**). Following inpatient admission for SARS-CoV-2 infections, 5.1% of patients required mechanical ventilation after a median 2.6 days (IQR: 0.8-6.5; **Table 2**). Median time to in-hospital death was 7.3 days (IQR: 3.7-13.3). Accounting for both in-hospital and out-of-hospital mortality, the 60-day risk of death after inpatient admission was 14.1%, with 11.3% of admitted patients dying without proceeding to mechanical ventilation, and 50.2% dying after initiating mechanical ventilation (**Table 4; Table S5**).

**Figure 3:**
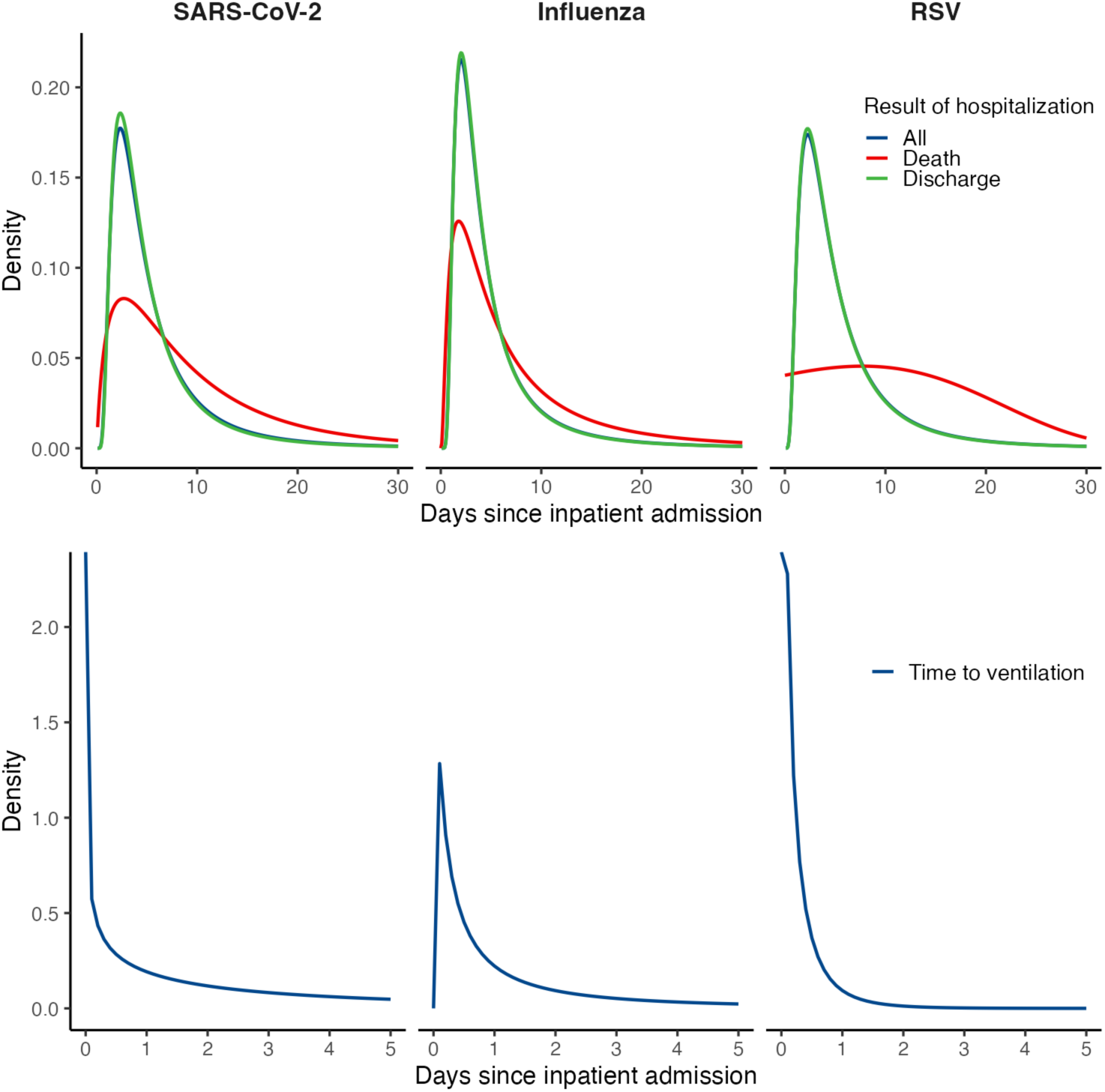
Durations of hospital stay. (*Top row*) We plot distributions from best-fitting models for durations of hospital stay, overall and stratified according to clinical outcome. (*Bottom row*) We plot distributions of time from inpatient admission to initiation of mechanical ventilation.

**Table 4:**
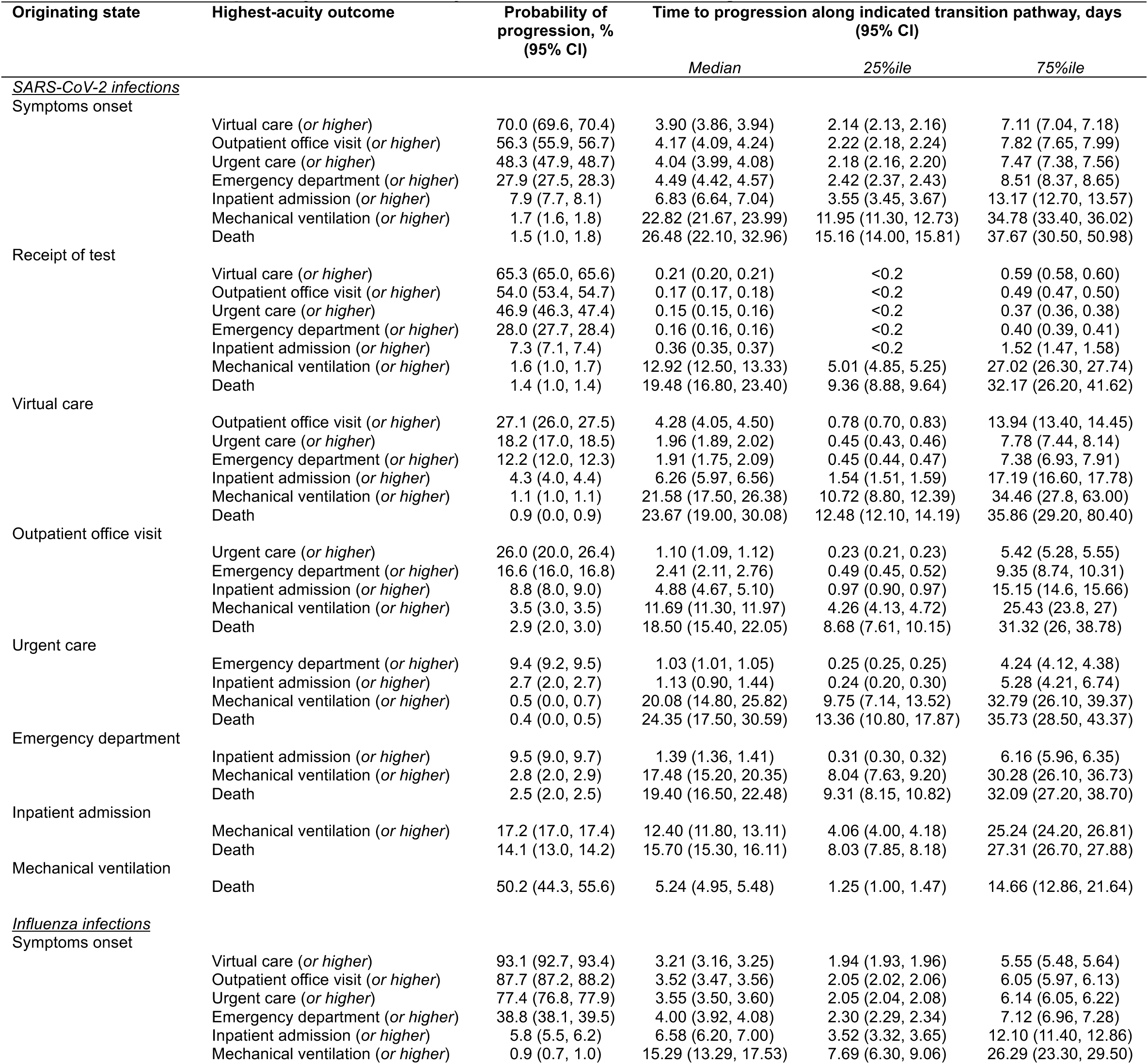
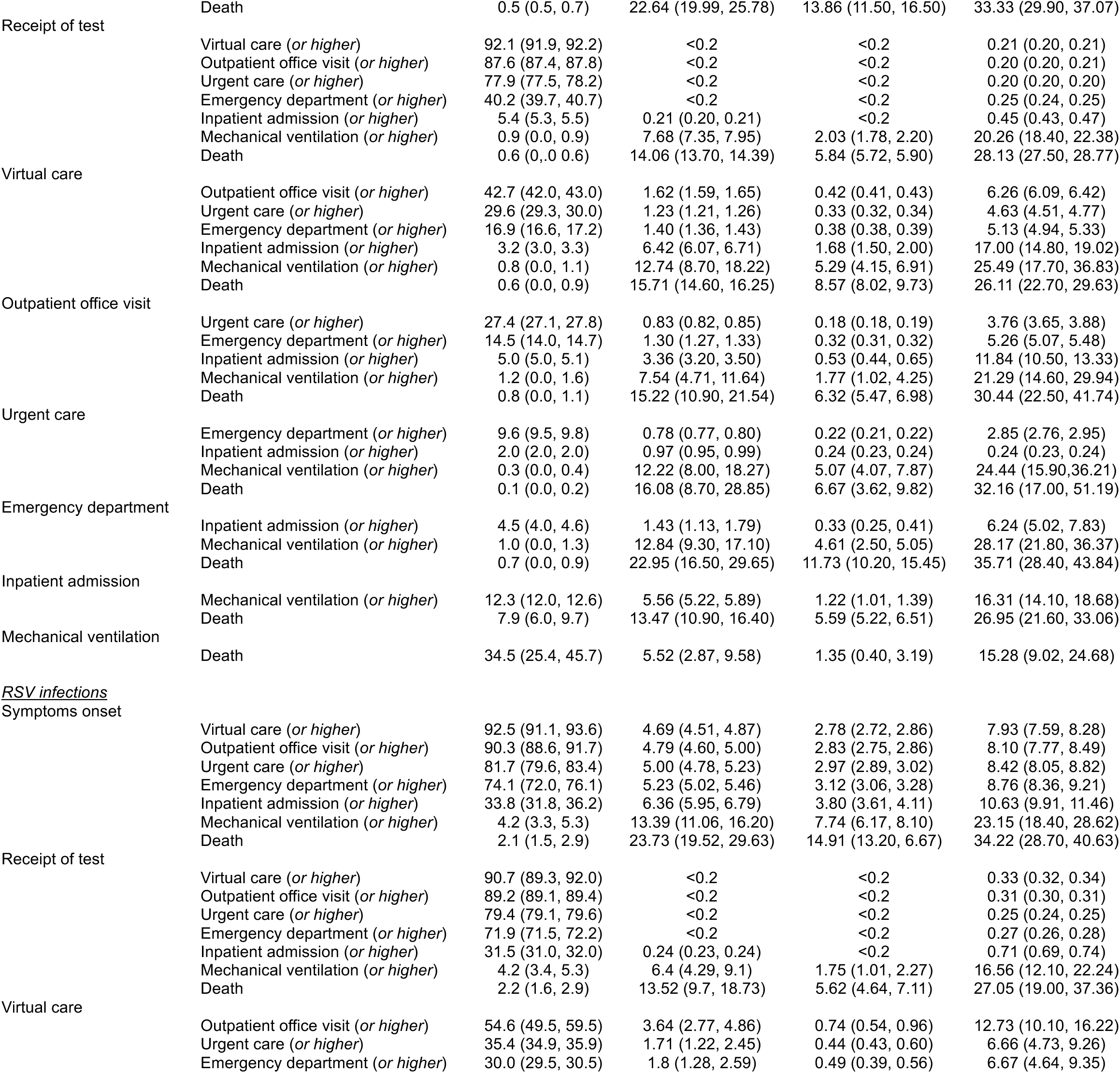
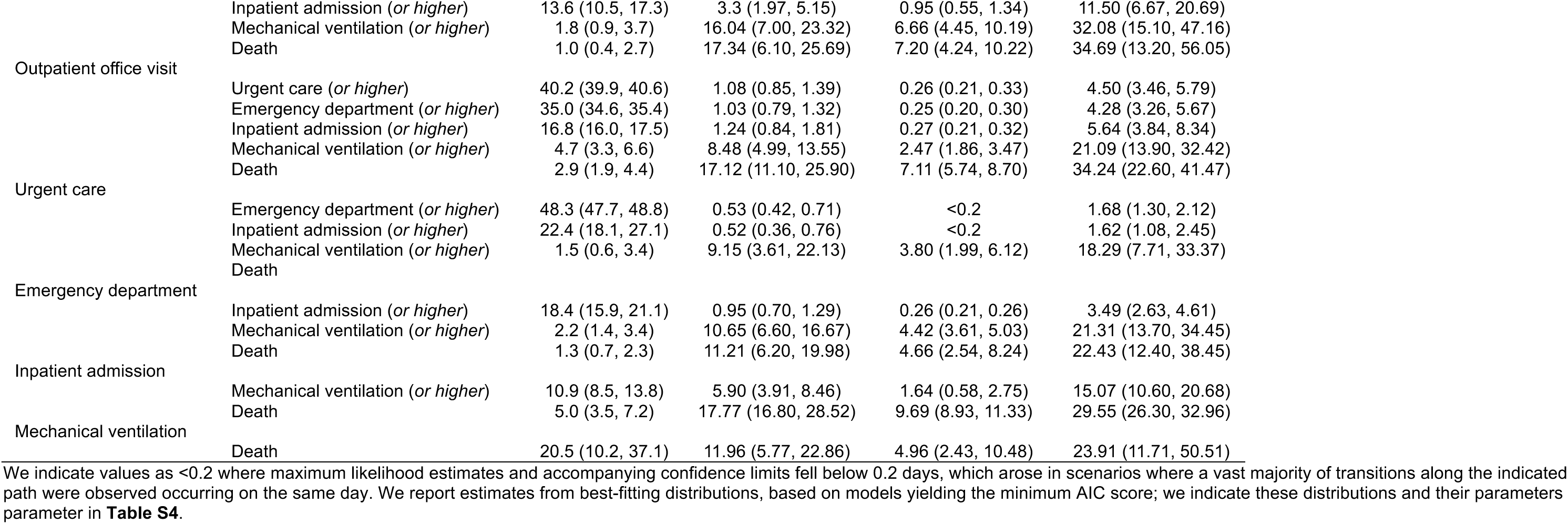
Care utilization pathways at each acuity threshold for each infecting virus.

### Care pathways for influenza and RSV

Median times from symptoms onset to testing were 3.4 and 5.6 days for influenza and RSV, respectively, corresponding to differences in the clinical care settings at which testing most frequently occurred for each pathogen (**Figure 2**). The first clinical encounter occurred in urgent care for 36.7% of influenza cases, in emergency departments for 28.8% of cases, and in hospital settings for 3.4%; a greater proportion of confirmed RSV infections (18.7%) were first intercepted in hospital settings (**Table 2**).

Median durations of hospital stay for influenza cases and RSV cases were 4.0 days (IQR: 2.3-6.8) and 4.3 days (IQR: 2.5-7.4), respectively (**Table S7**). The 60-day risk of death after hospital admission was 7.9% among influenza cases and 5.0% among RSV cases (**Table 4**). Median time to in-hospital death following admission was 5.2 days (IQR: 2.6-10.5) among influenza cases and 11.3 days (IQR: 5.8-17.6) among RSV cases. In the 60 days following an inpatient admission, median times from admission to death were 17.5 days (IQR: 4.1-31.4) and 18.7 days (9.2-30.0) for influenza and RSV cases, respectively, who did not require mechanical ventilation, while median times from initiation of mechanical ventilation to death were 5.5 days (1.4-15.3) and 12.0 days (5.0-23.9) for influenza and RSV cases, respectively (**Table S5**).

### Care requirements for all observed infections

For SARS-CoV-2 infections, median times from symptoms onset to receipt of care at or above the virtual care, outpatient physician office, urgent care, or emergency department thresholds were in the range of 3.9-4.5 days (**Table 3**). Overall, 7.9% of observed SARS-CoV-2 infections resulted in inpatient admission or death, occurring a median 6.8 days (IQR: 3.6-13.2) after symptoms onset. Progression to illness necessitating mechanical ventilation and death occurred markedly later in the course of illness (median 22.8 days [IQR: 12.0-34.8] and 26.2 days [IQR: 15.2-37.7] after symptoms onset, respectively) than initial inpatient admission.

**Table 3:** Observed proportions of cases attaining or exceeding each acuity threshold.

| Infecting virus | Acuity threshold | Probability of progression, % (95% CI) | Time to progression, days (95% CI) |  |  |
| --- | --- | --- | --- | --- | --- |
|  |  | Proportion, % (95% CI) | Median | 25%ile | 75%ile |
| SARS-CoV-2 | Virtual care (*or higher*) | 70.0 (69.6, 70.4) | 3.90 (3.86, 3.94) | 2.14 (2.13, 2.16) | 7.11 (7.04, 7.18) |
|  | Outpatient office visit (*or higher*) | 56.3 (55.9, 56.7) | 4.17 (4.09, 4.24) | 2.22 (2.18, 2.24) | 7.82 (7.65, 7.99) |
|  | Urgent care (*or higher*) | 48.3 (47.9, 48.7) | 4.04 (3.99, 4.08) | 2.18 (2.16, 2.20) | 7.47 (7.38, 7.56) |
|  | Emergency department (*or higher*) | 27.9 (27.5, 28.3) | 4.49 (4.42, 4.57) | 2.42 (2.37, 2.43) | 8.51 (8.37, 8.65) |
|  | Inpatient admission (*or higher*) | 7.9 (7.7, 8.1) | 6.83 (6.64, 7.04) | 3.55 (3.45, 3.67) | 13.17 (12.70, 13.57) |
|  | Mechanical ventilation (*or higher*) | 1.7 (1.6, 1.8) | 22.82 (21.67, 23.99) | 11.95 (11.30, 12.73) | 34.78 (33.40, 36.02) |
|  | Death | 1.3 (1.3, 1.4) | 26.20 (24.93, 27.56) | 15.16 (14.00, 15.81) | 37.67 (30.50, 50.98) |
| Influenza | Virtual care (*or higher*) | 93.1 (92.7, 93.4) | 3.21 (3.16, 3.25) | 1.94 (1.93, 1.96) | 5.55 (5.48, 5.64) |
|  | Outpatient office visit (*or higher*) | 87.7 (87.2, 88.2) | 3.52 (3.47, 3.56) | 2.05 (2.02, 2.06) | 6.05 (5.97, 6.13) |
|  | Urgent care (*or higher*) | 77.4 (76.8, 77.9) | 3.55 (3.50, 3.60) | 2.05 (2.04, 2.08) | 6.14 (6.05, 6.22) |
|  | Emergency department (*or higher*) | 38.8 (38.1, 39.5) | 4.00 (3.92, 4.08) | 2.30 (2.29, 2.34) | 7.12 (6.96, 7.28) |
|  | Inpatient admission (*or higher*) | 5.8 (5.5, 6.2) | 6.58 (6.20, 7.00) | 3.52 (3.32, 3.65) | 12.10 (11.40, 12.86) |
|  | Mechanical ventilation (*or higher*) | 0.9 (0.7, 1.0) | 15.29 (13.29, 17.53) | 7.69 (6.30, 9.06) | 26.29 (23.30, 29.50) |
|  | Death | 0.5 (0.5, 0.7) | 22.64 (19.99, 25.78) | 13.86 (11.50, 16.50) | 33.33 (29.90, 37.07) |
| RSV | Virtual care (*or higher*) | 92.5 (91.1, 93.6) | 4.69 (4.51, 4.87) | 2.78 (2.72, 2.86) | 7.93 (7.59, 8.28) |
|  | Outpatient office visit (*or higher*) | 90.3 (88.6, 91.7) | 4.79 (4.60, 5.00) | 2.83 (2.75, 2.86) | 8.10 (7.77, 8.49) |
|  | Urgent care (*or higher*) | 81.7 (79.6, 83.4) | 5.00 (4.78, 5.23) | 2.97 (2.89, 3.02) | 8.42 (8.05, 8.82) |
|  | Emergency department (*or higher*) | 74.1 (72.0, 76.1) | 5.23 (5.02, 5.46) | 3.12 (3.06, 3.28) | 8.76 (8.36, 9.21) |
|  | Inpatient admission (*or higher*) | 33.8 (31.8, 36.2) | 6.36 (5.95, 6.79) | 3.80 (3.61, 4.11) | 10.63 (9.91, 11.46) |
|  | Mechanical ventilation (*or higher*) | 4.2 (3.3, 5.3) | 13.39 (11.06, 16.20) | 7.74 (6.17, 8.10) | 23.15 (18.40, 28.62) |
|  | Death | 2.1 (1.5, 2.9) | 23.73 (19.52, 29.63) | 14.91 (13.20, 16.67) | 34.22 (28.70, 40.63) |

Nearly all influenza and RSV infections were linked to ARI diagnoses resulting from care appointments in any setting around the time of individuals’ first eligible positive test (93.1% and 92.5%, respectively; **Table 3**). Median times from symptoms onset to receipt of care at virtual, outpatient, urgent care, and emergency department or higher-acuity settings were 3.2, 3.5, 3.6, and 4.0 days, respectively, for influenza, and 4.7, 4.8, 5.0, and 5.2 days, respectively, for RSV (**Figure 2**). Median times from symptoms onset to inpatient admission, mechanical ventilation, and death were 6.4-6.6 days, 13.4-15.3 days, and 22.6-23.7 days, respectively.

### Associations of care trajectories with individual characteristics

The proportion of cases receiving care at each acuity level increased with older age for all infections; age differences were most pronounced for high-acuity outcomes (e.g., inpatient admission, mechanical ventilation, and mortality; **Figure 4; Table S8**; **Figure S1**; **Figure S2**). Median times from symptoms onset to receipt of care at or above the level of outpatient office visits increased with older age, spanning a difference of ∼1 day between the ≤17 year and ≥90 year age groups for all three viral infections (3.3 vs. 4.7 days for SARS-CoV-2 infections, 3.2 vs. 4.4 days for influenza infections, and 4.0 vs. 4.8 days for RSV infections), although these differences across ages in times to event were attenuated for higher-acuity outcomes. Individuals with greater numbers of comorbid conditions also had higher chances of receiving care at each level of acuity and longer median times to presentation, for each virus (**Table S11**).

**Figure 4:**
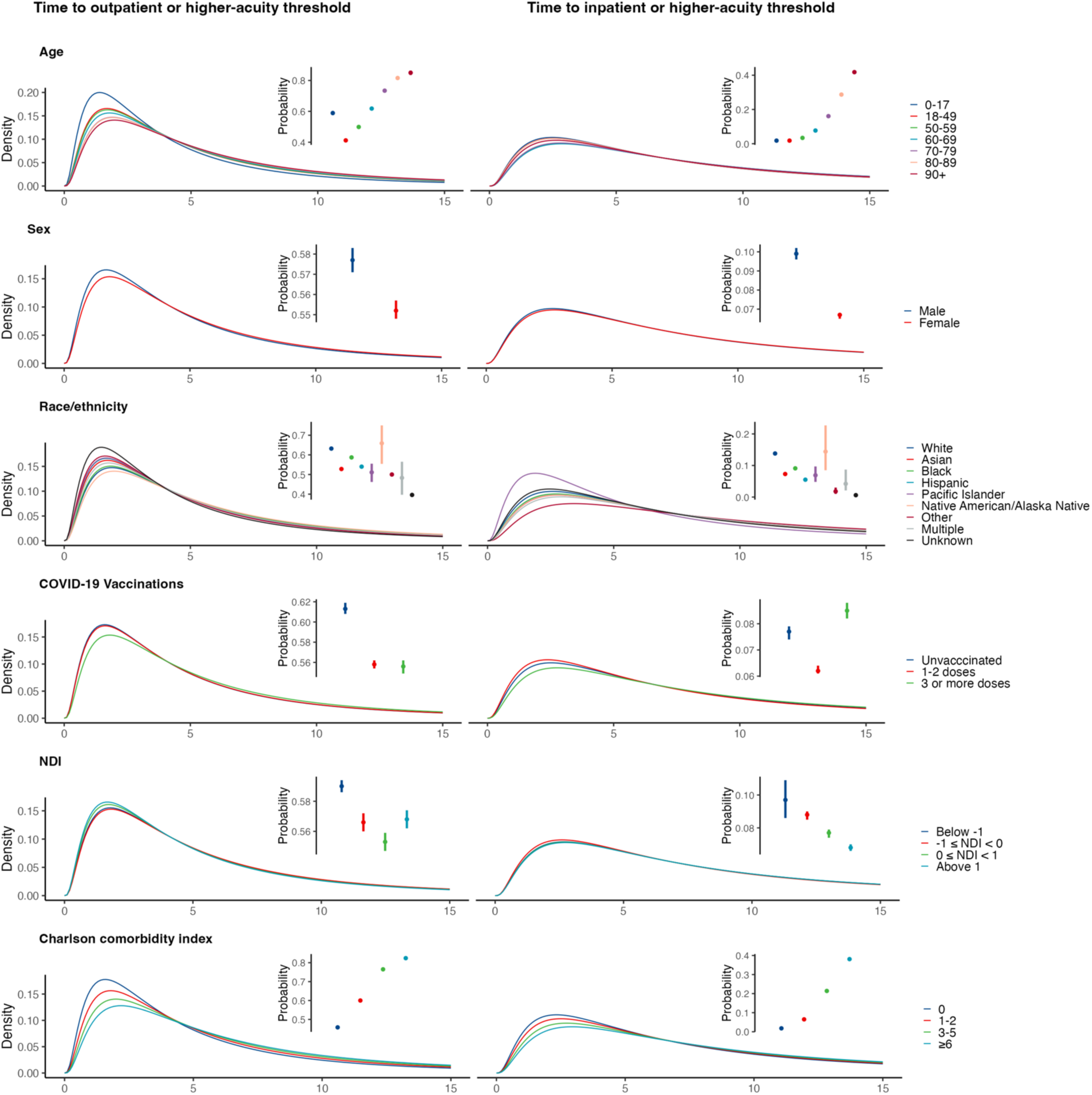
Probabilities and time-to-event distributions of reaching outpatient and inpatient acuity thresholds from symptom onset for illness associated with a SARS-CoV-2 infection across demographic subgroups. We illustrate best-fitting distributions for times to the indicated events from symptom onset. Probabilities of reaching acuity thresholds are available in **Tables S8**-**S12**. We present similar figures for influenza and RSV (**Figures S1** and **S2**).

Whereas a greater proportion of males than females with SARS-CoV-2 infection experienced high-acuity outcomes (e.g., 9.9% vs. 6.7% with inpatient admission or higher-acuity outcomes, 2.1% vs. 1.1% mortality; **Table S9**), this pattern was less clearly apparent for influenza cases and was reversed for RSV. Times to each outcome were similar for male and female cases with each infection. With regard to individuals’ vaccination status and neighborhood deprivation index values, we did not identify patterns across pathogens or across outcomes with respect to any subgroup experiencing consistently higher or lower likelihood of progression, or consistently longer or shorter times to progression (**Table S10**; **Table S12**).

The probability of in-hospital mortality for SARS-CoV-2 infections was higher among older adults compared to younger adults (11.0% at ages ≥90 years versus 2.1% in 18-49 year age group; **Table S13**). There were also significant differences across Charlson comorbidity subgroups, with 9.0% of cases with a score ≥6 experiencing in-hospital mortality associated with SARS-CoV-2 infection in comparison to 3.9% mortality among cases with a score of 0. This trend was also apparent in influenza infections (7.2% vs. 1.7%), although not in RSV infections (**Table S13)**.

Median durations of hospital stay were similar across groups for each infection. Males had longer median times to in-hospital mortality than females for SARS-CoV-2 infections (8.5 vs. 6.8 days), but shorter times to mortality for influenza and RSV infections (4.9 vs. 5.7 days and 8.1 vs. 16.1 days, respectively; **Table S13**). We did not observe differences across subgroups with respect to vaccination status, race or ethnicity, or neighborhood deprivation index in the probability of in-hospital mortality or length of hospital stays.

### Github repository

In addition to the descriptive supplementary materials associated with this manuscript, we have created a Github repository containing parameter estimates for all analyses described (https://github.com/ntparker3/Resp_params). The repository includes four files for each pathogen (SARS-CoV-2, influenza, and RSV), contents of which are listed below:

1. Parameterized distributions and summary statistics of the proximal progression event occurring from each originating state (“first event”);
2. Parameterized distributions and summary statistics for individuals’ to risk of progression to or above each acuity threshold, from each originating state (“event or worse”);
3. Probabilities of progression to or above each acuity threshold, from each originating state, across subgroups of the specified individual-level covariates (“event or worse covariates”); and
4. Location parameters and median times to event for progression to outpatient (or higher-acuity) and inpatient (or higher-acuity) thresholds, from each originating state, across subgroups of the specified individual-level covariates (“covariate rates”).

## DISCUSSION

Our analysis provides estimates of transition rates and probabilities for healthcare utilization due to the progression of ARIs associated with SARS-CoV-2, influenza, and RSV infections. These outputs aim to inform models anticipating resource needs for healthcare systems and public health stakeholders, drawing on real-world observations within a US managed care setting. In addition to presenting aggregated results for all cases infected with SARS-CoV-2, influenza, and RSV, we present stratified results for differing subgroups for which models may aim to generate predictions; these encompassed patient demographics (age, sex, race/ethnicity), comorbidity burden, prior vaccination, and community-level socioeconomic disadvantage. We identified the strongest evidence of differences in progression risk and times-to-event across age groups and comorbidity profiles. Our outputs fill frequently-described gaps in the data needed for application of viral respiratory infection models (17–19) and may inform future forecasting efforts tailored to US healthcare contexts (20,21).

Previous studies have reported widely varying estimates of times from symptoms onset to hospital admission for COVID-19 (22–25) and the duration of hospital stay among COVID-19 patients (7,8,26), with both parameters differing across settings and over time within settings in association with evolving clinical practices. Whereas numerous studies have monitored patients hospitalized with each virus (27,28), fewer have tracked outcomes longitudinally from early points in the disease course such as symptoms onset or receipt of care in virtual or ambulatory facilities. Within our study, only 4.7% of COVID-19 cases (7.9% of all COVID-19 cases who received care in any setting) were first intercepted at the point of hospital admission, while among influenza and RSV cases, 3.4% and 18.7%, respectively, were first seen in inpatient settings. These circumstances suggest that projecting outcomes among individuals receiving care in lower-acuity settings may help to refine forecasts of higher-acuity clinical care needs.

Application of our estimates to forecasting models requires several assumptions or considerations. First, we frame consecutive transitions between states as memoryless, consistent with modeling approaches where estimates from these analyses may be applied (e.g., Markov chain next-state transitions as well as cumulative probabilities of attaining each state overall and from preceding states). Second, our analyses are subset to individuals who ultimately received care including diagnostic testing: events preceding testing among individuals included in these analyses, particularly in low-acuity care settings, may not represent care utilization pathways among individuals who were ultimately never tested—a problem related to previously described biases affecting interval distribution estimation (29–31). Furthermore, testing for influenza and RSV was more strictly limited to individuals who received care in outpatient or inpatient facilities, whereas SARS-CoV-2 tests were widely available across all care settings. In particular, RSV testing in adults is restricted to inpatient settings, with the majority of testing, and subsequent recorded cases, being conducted in children. Thus, differences in the overall proportions of SARS-CoV-2 infections, influenza infections, and RSV infections attaining each acuity threshold should not be interpreted as differences in the severity of disease caused by each infection. Last, associations of the studied covariates with the proportions of cases experiencing each outcome and with times-to-event should not be interpreted as causal. In some instances, observed patterns reflect previously reported independent associations, such as associations of older age and the presence of comorbidities with severe disease outcomes (32).

However, other findings, such as the lack of association of prior vaccination with protection against severe outcomes, echo previous evidence of higher uptake of vaccines against COVID-19, seasonal influenza, and RSV among individuals at greatest risk (33–35). These analyses aim to inform prediction even if they lack direct causal interpretation.

Our analysis has at least 6 limitations. First, KPSC represents a single healthcare system. While strengths include the integration of care delivery and data capture across outpatient and inpatient settings, the large enrollee population, and its racial/ethnic and socioeconomic diversity (13), it remains important to note that care utilization and delivery pathways may not be generalizable to all settings. Estimation of similar parameters in other US populations remains an important objective. Second, aiming to enable the broad application of our estimates, we fit parametric distributions to times-to-event that may not perfectly represent underlying processes. For this reason, we supply best-fitting parameter estimates for 6 different distributions of all times-to-event, and report which distributions provided the best fit to data based on AIC values.

Third, major SARS-CoV-2 variants (e.g., XBB, BA.2.86/JN.1, and KP.2) and seasonal influenza lineages (e.g., A(H1N1)pdm09, A(H3N2), B(Victoria)) circulating during the study period may not generalize to lineages circulating during future years. Fourth, the reliability of self-reported and physician-recorded symptoms onset dates may be imperfect, as signified by patterns such as heaping of times from symptoms onset around 7 days and 14 days before testing and other healthcare encounters (36). Fifth, recovery was not explicitly recorded as a clinical outcome, necessitating censoring of observation periods without future healthcare encounters. Last, our analyses preceded the widespread implementation of RSV vaccines among pregnant mothers and older adults, which may alter RSV-related healthcare utilization in future seasons for population groups at the highest risk of severe disease.

These limitations notwithstanding, our analyses provide a useful entry point for modeling real-world trajectories of healthcare needs associated with SARS-CoV-2, influenza, and RSV infections. Extensive ARI-associated healthcare utilization in virtual, outpatient, and urgent care settings among persons ultimately hospitalized suggests monitoring of lower-acuity healthcare utilization may help to inform near-term hospital capacity requirements. Incorporating data on lower-acuity care delivery settings into public health surveillance and reporting thus merits consideration. Similar analyses in other geographic settings or other healthcare systems, and continued updating of parameters we report to accommodate changes in viral epidemiology or healthcare delivery practices, may improve the reliability of forecasting models for SARS-CoV-2, influenza, and RSV.

## Data Availability

In addition to the descriptive supplementary materials associated with this manuscript, we have created a Github repository containing parameter estimates for all analyses described (https://github.com/ntparker3/Resp_params). The raw data used to develop models in this manuscript include protected health information (e.g. dates of diagnoses and testing) that cannot be shared openly without appropriate human subjects approval and data use agreements. Kaiser Permanente Southern California (KPSC) institutional policy requires a data transfer agreement to be executed between KPSC and the individual recipient entity prior to transmittal of patient-level data outside KPSC. Requests for data can be addressed to the Central Business Office of the Department of Research and Evaluation (contact via).

## ACKNOWLEDGMENTS

This work was funded by the Center for Forecasting and Outbreak Analytics of the US Centers for Disease Control & Prevention via the Insight Net cooperative agreement (CDC-RFA-FT-23-0069 to SYT and JAL). Its contents are solely the responsibility of the authors and do not necessarily represent the official views of the Centers for Disease Control and Prevention.

## AUTHOR CONTRIBUTIONS

**Conceptualization:** SYT, JAL

**Data curation:** VH, FX

**Formal analysis:** NTP

**Funding acquisition:** SYT, JAL

**Investigation:** NTP, VH, GSD, FX

**Methodology:** NTP, VH, FX, IRB, SYT, JAL

**Project administration:** MP, IAR

**Resources/software:** NTP

**Supervision:** NFM, IRB, SYT, JAL

**Visualization:** NTP

**Writing—original draft:** NTP, JAL

**Writing—review & editing:** VH, GSD, MP, IAR, FX, NFM, IRB, SYT

---

**Table S1:**
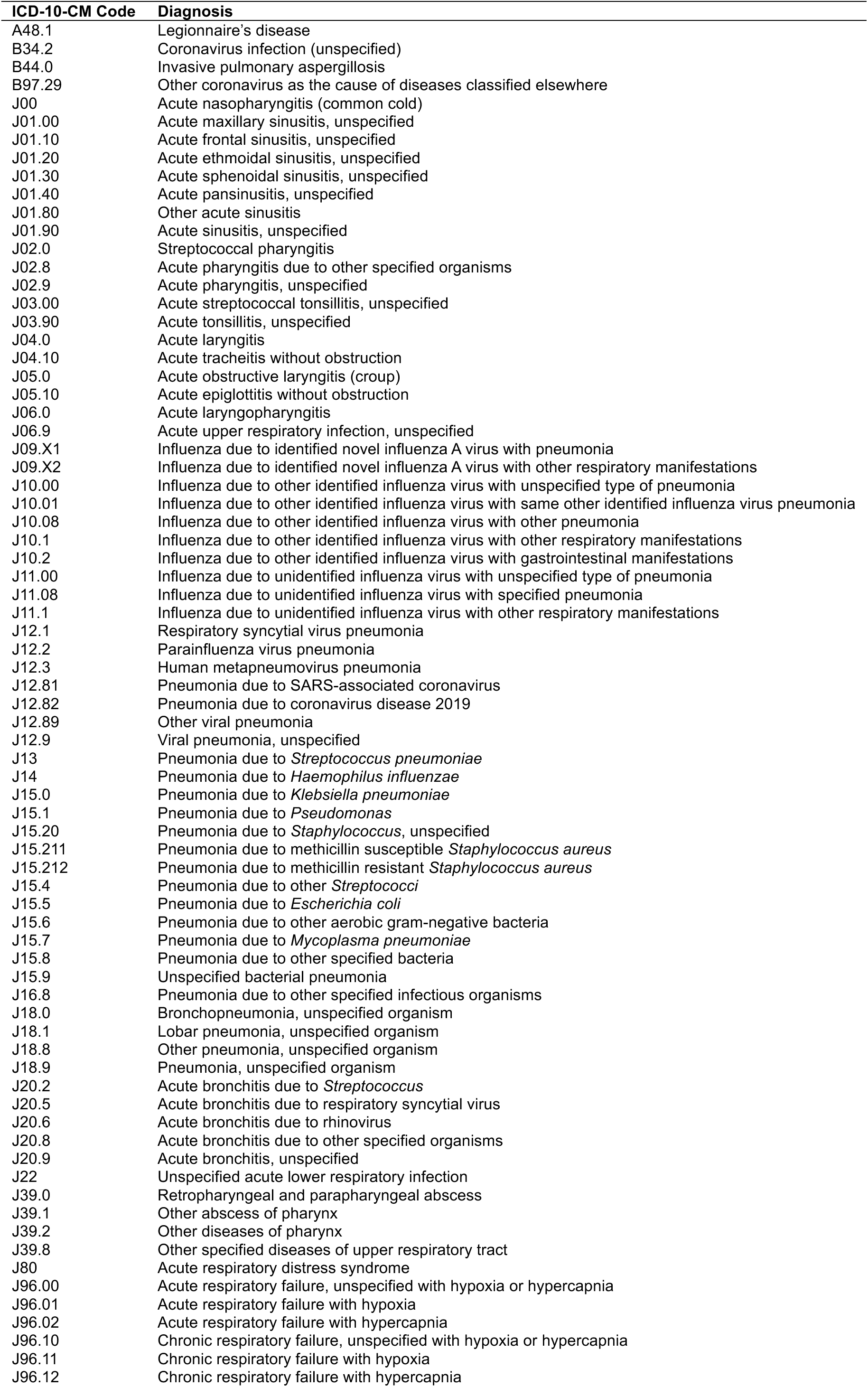
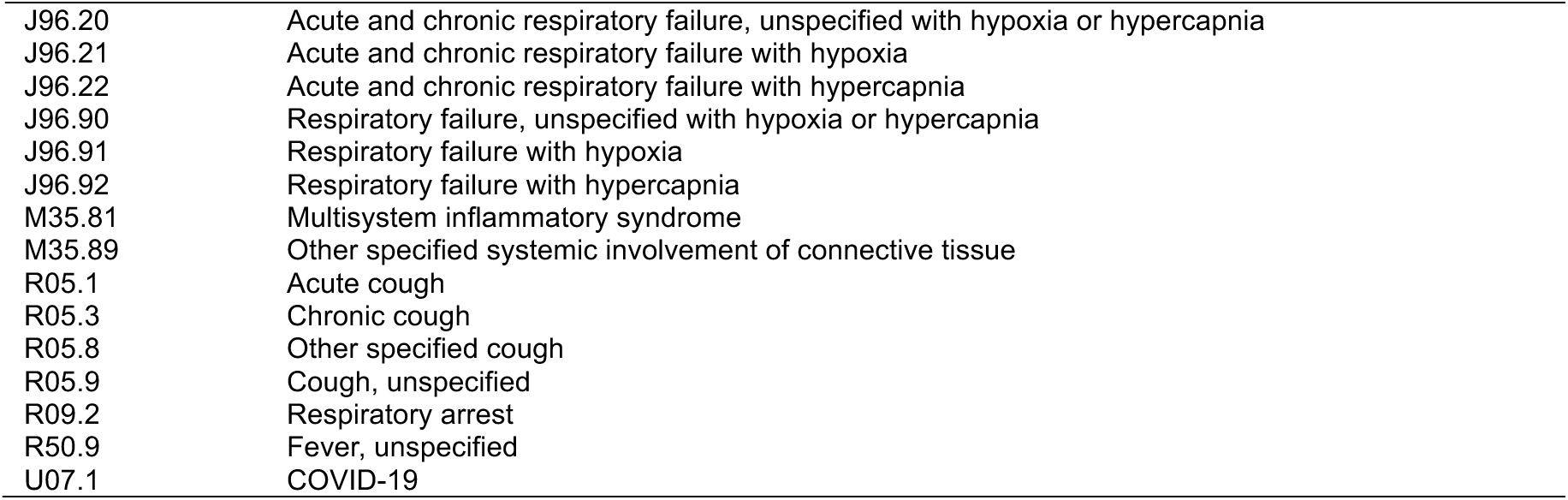
Acute respiratory infection diagnosis codes.

| ICD-10-CM Code | Diagnosis |
| --- | --- |
| A48.1 | Legionnaire's disease |
| B34.2 | Coronavirus infection (unspecified) |
| B44.0 | Invasive pulmonary aspergillosis |
| B97.29 | Other coronavirus as the cause of diseases classified elsewhere |
| J00 | Acute nasopharyngitis (common cold) |
| J01.00 | Acute maxillary sinusitis, unspecified |
| J01.10 | Acute frontal sinusitis, unspecified |
| J01.20 | Acute ethmoidal sinusitis, unspecified |
| J01.30 | Acute sphenoidal sinusitis, unspecified |
| J01.40 | Acute pansinusitis, unspecified |
| J01.80 | Other acute sinusitis |
| J01.90 | Acute sinusitis, unspecified |
| J02.0 | Streptococcal pharyngitis |
| J02.8 | Acute pharyngitis due to other specified organisms |
| J02.9 | Acute pharyngitis, unspecified |
| J03.00 | Acute streptococcal tonsillitis, unspecified |
| J03.90 | Acute tonsillitis, unspecified |
| J04.0 | Acute laryngitis |
| J04.10 | Acute tracheitis without obstruction |
| J05.0 | Acute obstructive laryngitis (croup) |
| J05.10 | Acute epiglottitis without obstruction |
| J06.0 | Acute laryngopharyngitis |
| J06.9 | Acute upper respiratory infection, unspecified |
| J09.X1 | Influenza due to identified novel influenza A virus with pneumonia |
| J09.X2 | Influenza due to identified novel influenza A virus with other respiratory manifestations |
| J10.00 | Influenza due to other identified influenza virus with unspecified type of pneumonia |
| J10.01 | Influenza due to other identified influenza virus with same other identified influenza virus pneumonia |
| J10.08 | Influenza due to other identified influenza virus with other pneumonia |
| J10.1 | Influenza due to other identified influenza virus with other respiratory manifestations |
| J10.2 | Influenza due to other identified influenza virus with gastrointestinal manifestations |
| J11.00 | Influenza due to unidentified influenza virus with unspecified type of pneumonia |
| J11.08 | Influenza due to unidentified influenza virus with specified pneumonia |
| J11.1 | Influenza due to unidentified influenza virus with other respiratory manifestations |
| J12.1 | Respiratory syncytial virus pneumonia |
| J12.2 | Parainfluenza virus pneumonia |
| J12.3 | Human metapneumovirus pneumonia |
| J12.81 | Pneumonia due to SARS-associated coronavirus |
| J12.82 | Pneumonia due to coronavirus disease 2019 |
| J12.89 | Other viral pneumonia |
| J12.9 | Viral pneumonia, unspecified |
| J13 | Pneumonia due to *Streptococcus pneumoniae* |
| J14 | Pneumonia due to *Haemophilus influenzae* |
| J15.0 | Pneumonia due to *Klebsiella pneumoniae* |
| J15.1 | Pneumonia due to *Pseudomonas* |
| J15.20 | Pneumonia due to *Staphylococcus*, unspecified |
| J15.211 | Pneumonia due to methicillin susceptible *Staphylococcus aureus* |
| J15.212 | Pneumonia due to methicillin resistant *Staphylococcus aureus* |
| J15.4 | Pneumonia due to other *Streptococci* |
| J15.5 | Pneumonia due to *Escherichia coli* |
| J15.6 | Pneumonia due to other aerobic gram-negative bacteria |
| J15.7 | Pneumonia due to *Mycoplasma pneumoniae* |
| J15.8 | Pneumonia due to other specified bacteria |
| J15.9 | Unspecified bacterial pneumonia |
| J16.8 | Pneumonia due to other specified infectious organisms |
| J18.0 | Bronchopneumonia, unspecified organism |
| J18.1 | Lobar pneumonia, unspecified organism |
| J18.8 | Other pneumonia, unspecified organism |
| J18.9 | Pneumonia, unspecified organism |
| J20.2 | Acute bronchitis due to *Streptococcus* |
| J20.5 | Acute bronchitis due to respiratory syncytial virus |
| J20.6 | Acute bronchitis due to rhinovirus |
| J20.8 | Acute bronchitis due to other specified organisms |
| J20.9 | Acute bronchitis, unspecified |
| J22 | Unspecified acute lower respiratory infection |
| J39.0 | Retropharyngeal and parapharyngeal abscess |
| J39.1 | Other abscess of pharynx |
| J39.2 | Other diseases of pharynx |
| J39.8 | Other specified diseases of upper respiratory tract |
| J80 | Acute respiratory distress syndrome |
| J96.00 | Acute respiratory failure, unspecified with hypoxia or hypercapnia |
| J96.01 | Acute respiratory failure with hypoxia |
| J96.02 | Acute respiratory failure with hypercapnia |
| J96.10 | Chronic respiratory failure, unspecified with hypoxia or hypercapnia |
| J96.11 | Chronic respiratory failure with hypoxia |
| J96.12 | Chronic respiratory failure with hypercapnia |
| J96.20 | Acute and chronic respiratory failure, unspecified with hypoxia or hypercapnia |
| J96.21 | Acute and chronic respiratory failure with hypoxia |
| J96.22 | Acute and chronic respiratory failure with hypercapnia |
| J96.90 | Respiratory failure, unspecified with hypoxia or hypercapnia |
| J96.91 | Respiratory failure with hypoxia |
| J96.92 | Respiratory failure with hypercapnia |
| M35.81 | Multisystem inflammatory syndrome |
| M35.89 | Other specified systemic involvement of connective tissue |
| R05.1 | Acute cough |
| R05.3 | Chronic cough |
| R05.8 | Other specified cough |
| R05.9 | Cough, unspecified |
| R09.2 | Respiratory arrest |
| R50.9 | Fever, unspecified |
| U07.1 | COVID-19 |

**Table S2:** Individual characteristics by infecting virus and severity threshold.

| Characteristic |  | Cases, according to infecting virus and severity threshold, n (%) |  |  |  |  |  |
| --- | --- | --- | --- | --- | --- | --- | --- |
|  |  | Infections receiving ARI diagnoses in any setting |  |  | Infections resulting in inpatient admission |  |  |
|  |  | SARS-CoV-2 | Influenza | RSV | SARS-CoV-2 | Influenza | RSV |
|  |  | N=39,231 | N=18,689 | N=1,550 | N=4,772 | N=1,172 | N=558 |
| Age, years | 0-17 | 2,863 (7.3) | 5,563 (29.8) | 842 (54.3) | 74 (1.6) | 85 (7.3) | 129 (23.1) |
|  | 18-49 | 12,674 (32.3) | 6,436 (34.4) | 65 (4.2) | 427 (8.9) | 180 (15.4) | 28 (5.0) |
|  | 50-59 | 5,961 (15.2) | 2,356 (12.6) | 80 (5.2) | 334 (7.0) | 105 (9.0) | 47 (8.4) |
|  | 60-69 | 6,380 (16.3) | 2,051 (11.0) | 138 (8.9) | 693 (14.5) | 208 (17.7) | 78 (14.0) |
|  | 70-79 | 6,381 (16.3) | 1,525 (8.2) | 167 (10.8) | 1,355 (28.4) | 317 (27.0) | 99 (17.7) |
|  | 80-89 | 3,937 (10.0) | 621 (3.3) | 187 (12.1) | 1,370 (28.7) | 210 (17.9) | 127 (22.8) |
|  | ≥90 | 1,035 (2.6) | 137 (0.7) | 71 (4.6) | 519 (10.9) | 67 (5.7) | 50 (9.0) |
| Sex | Male | 15,823 (40.3) | 8,450 (45.2) | 716 (46.2) | 2,377 (49.8) | 534 (45.6) | 234 (41.9) |
|  | Female | 23,408 (59.7) | 10,239 (54.8) | 834 (53.8) | 2,395 (50.2) | 638 (54.4) | 324 (58.1) |
| Race/ethnicity | White, non-Hispanic | 10,223 (26.1) | 4,154 (22.2) | 451 (29.1) | 2,020 (42.3) | 404 (34.5) | 206 (36.9) |
|  | Asian, non-Hispanic | 4,403 (11.2) | 1,990 (10.6) | 190 (12.3) | 490 (10.3) | 142 (12.1) | 66 (11.8) |
|  | Black, non-Hispanic | 4,494 (11.5) | 1,783 (9.5) | 154 (9.9) | 612 (12.8) | 135 (11.5) | 56 (10.0) |
|  | Hispanic (any race) | 18,282 (46.6) | 9,732 (52.1) | 705 (45.5) | 1,564 (32.8) | 472 (40.3) | 214 (38.4) |
|  | Native American/Alaska native | 87 (0.2) | 27 (0.1) | 3 (0.2) | 16 (0.3) | 4 (0.3) | 3 (0.5) |
|  | Other | 528 (1.3) | 310 (1.7) | 11 (0.7) | 15 (0.3) | 3 (0.3) | 2 (0.4) |
|  | Multiple | 128 (0.3) | 90 (0.5) | 8 (0.5) | 10 (0.2) | 1 (0.1) | 1 (0.2) |
|  | Pacific Islander | 303 (0.8) | 131 (0.7) | 12 (0.8) | 37 (0.8) | 9 (0.8) | 8 (1.4) |
|  | Unknown | 783 (2.0) | 472 (2.5) | 16 (1.0) | 8 (0.2) | 2 (0.2) | 2 (0.4) |
| Insurance source | Commercial | 18,057 (46.0) | 10,304 (55.1) | 634 (40.9) | 823 (17.2) | 314 (26.8) | 143 (25.6) |
|  | Medicaid | 4,437 (11.3) | 2,651 (14.2) | 279 (18.0) | 512 (10.7) | 158 (13.5) | 92 (16.5) |
|  | Medicare | 10,665 (27.2) | 2,284 (12.2) | 381 (24.6) | 2,898 (60.7) | 532 (45.4) | 241 (43.2) |
|  | Pre-paid plans | 945 (2.4) | 542 (2.9) | 30 (1.9) | 46 (1.0) | 20 (1.7) | 10 (1.8) |
|  | Other | 473 (1.2) | 154 (0.8) | 14 (0.9) | 97 (2.0) | 18 (1.5) | 5 (0.9) |
|  | Unknown | 4,654 (11.9) | 2,754 (14.7) | 212 (13.7) | 396 (8.3) | 130 (11.1) | 67 (12.0) |
| Vaccinations | 1-2 COVID-19 vaccine doses | 7,263 (18.5) | 4,285 (22.9) | 147 (9.5) | 706 (14.8) | 200 (17.1) | 62 (11.1) |
|  | 3+ COVID-19 vaccine doses | 27,174 (69.3) | 9,336 (50.0) | 682 (44.0) | 3,542 (74.2) | 797 (68.0) | 371 (66.5) |
| Flu Vaccination | Seasonal influenza vaccine received | 11,347 (28.9) | 8,376 (44.8) | 349 (22.5) | 744 (15.6) | 275 (23.5) | 468 (16.1) |
|  | RSV vaccine received | 339 (0.9) | 179 (1.0) | 26 (1.7) | 68 (1.4) | 27 (2.3) | 12 (2.2) |
| Neighborhood deprivation index | NDI < -1 | 1,775 (4.4) | 837 (4.5) | 68 (4.4) | 253 (5.3) | 46 (3.9) | 28 (5.0) |
|  | -1 ≤ NDI < 0 | 13,331 (32.9) | 5,821 (31.1) | 499 (32.2) | 1,704 (35.7) | 387 (33.0) | 167 (29.9) |
|  | 0 ≤ NDI < 1 | 13,943 (34.4) | 6,520 (34.9) | 535 (34.5) | 1,610 (33.7) | 427 (36.4) | 205 (36.7) |
|  | NDI > 1 | 7,583 (18.7) | 3,681 (19.7) | 262 (16.9) | 765 (16.0) | 212 (18.1) | 92 (16.5) |
|  | Unknown | 3,904 (9.6) | 1,830 (9.8) | 186 (12.0) | 440 (9.2) | 100 (8.5) | 66 (11.8) |
| Charlson comorbidity index | 0 | 19,220 (47.4) | 11,832 (63.3) | 763 (49.2) | 602 (12.6) | 226 (19.3) | 120 (21.5) |
|  | 1-2 | 11,905 (29.4) | 4,761 (25.5) | 354 (22.8) | 1,090 (22.8) | 342 (29.2) | 142 (25.4) |
|  | 3-5 | 6,042 (14.9) | 1,372 (7.3) | 243 (15.7) | 1,582 (33.2) | 332 (28.3) | 154 (27.6) |
|  | 6+ | 3,369 (8.3) | 724 (3.9) | 190 (12.3) | 1,498 (31.4) | 272 (23.2) | 142 (25.4) |

ARI: Acute respiratory infection. Diagnosis codes are listed in **Table S1**.

**Table S3:**
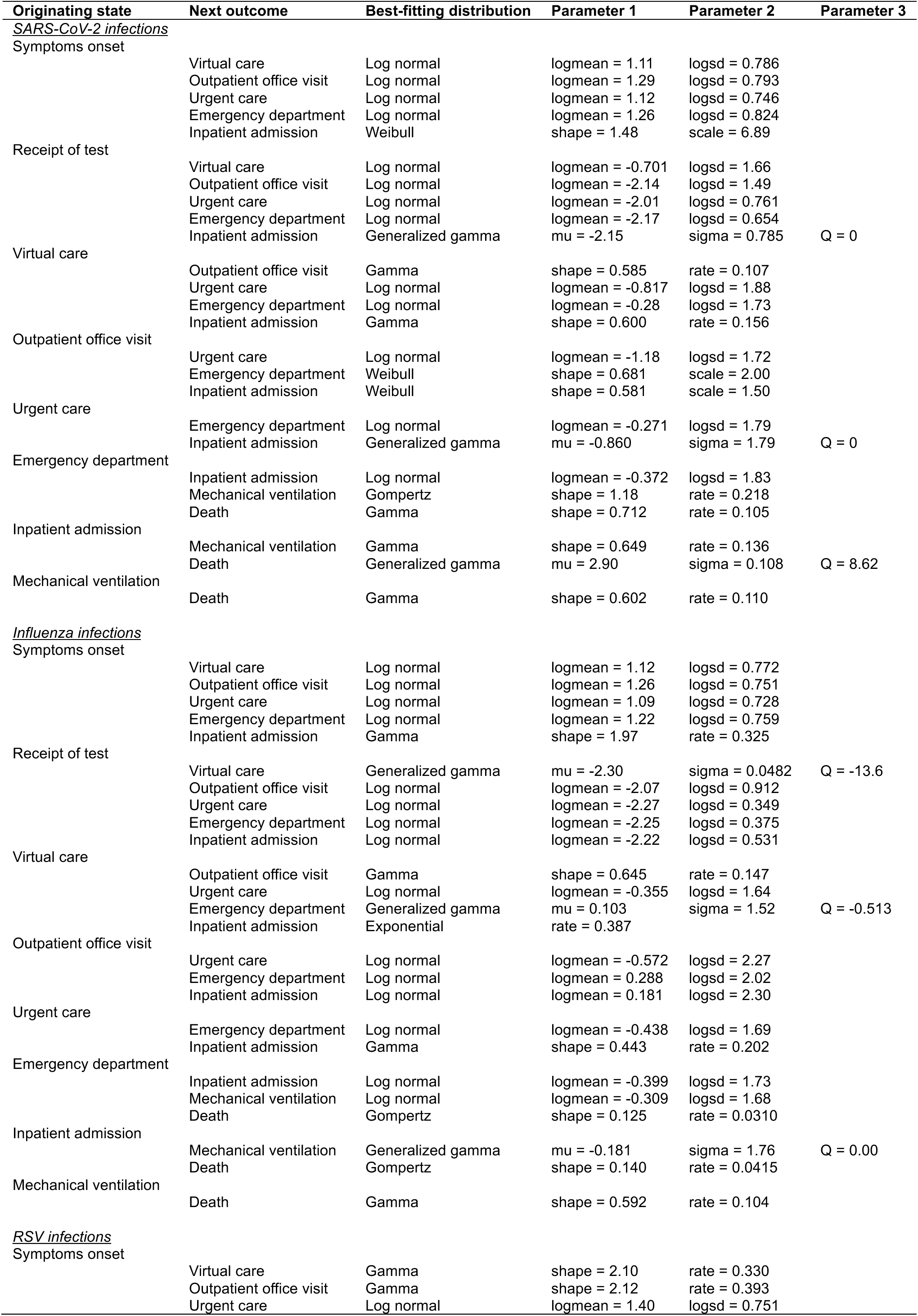
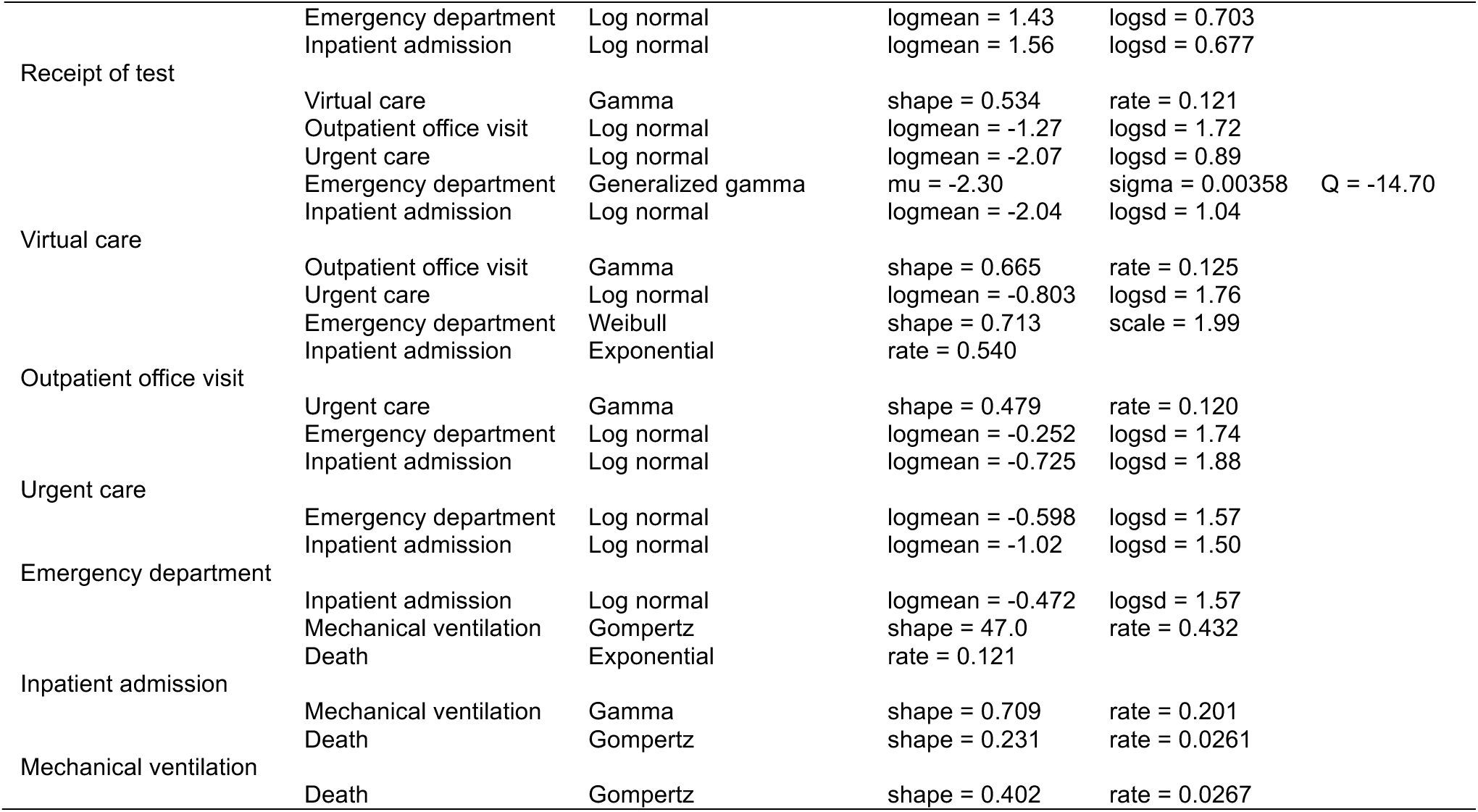
Best-fitting distributions for care utilization pathways for all infections using a 20 day follow-up period.

**Table S4:**
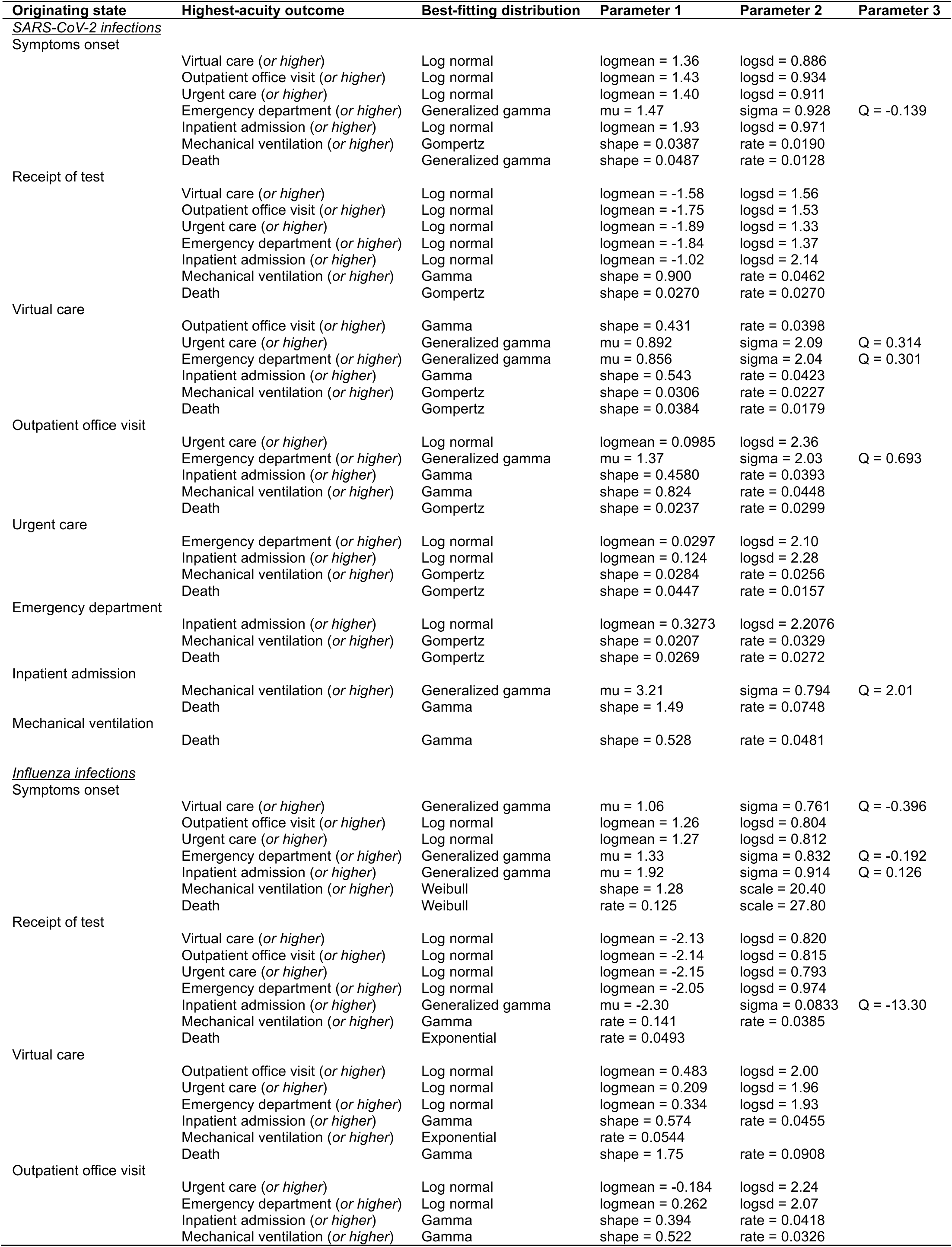
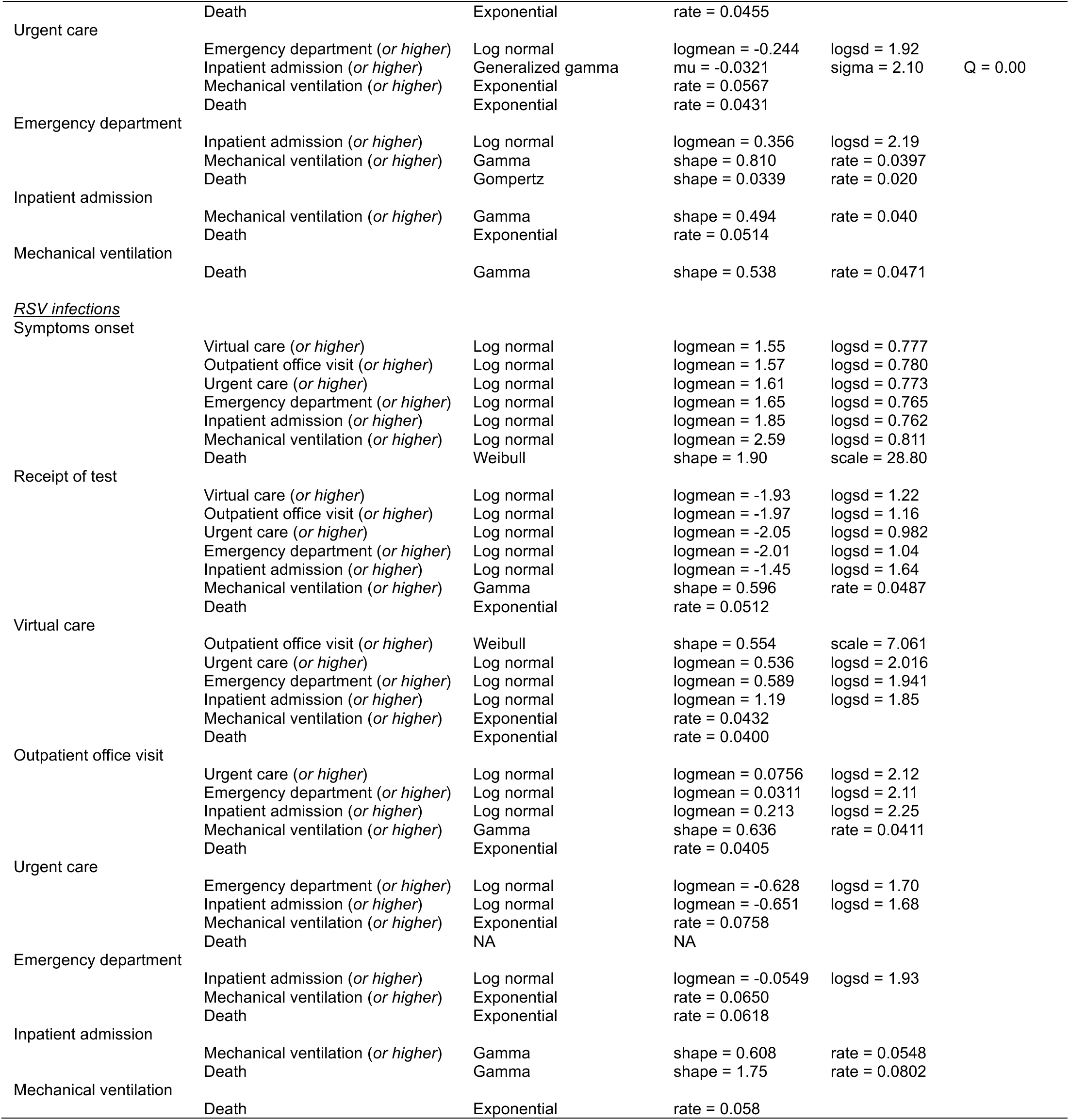
Best-fitting distributions for care utilization pathways at each acuity threshold for each infection using a 60 day follow-up period.

**Table S5:**
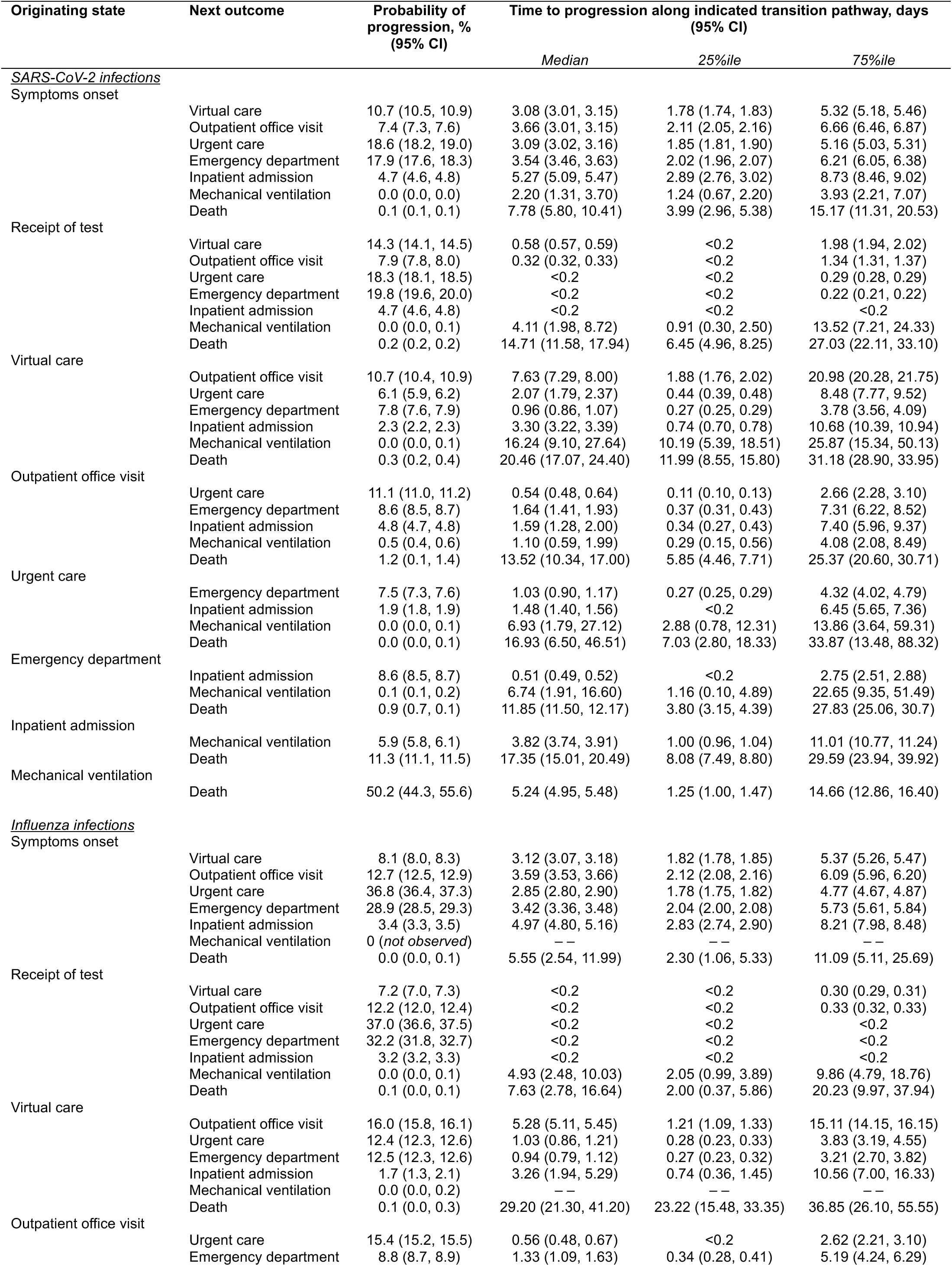
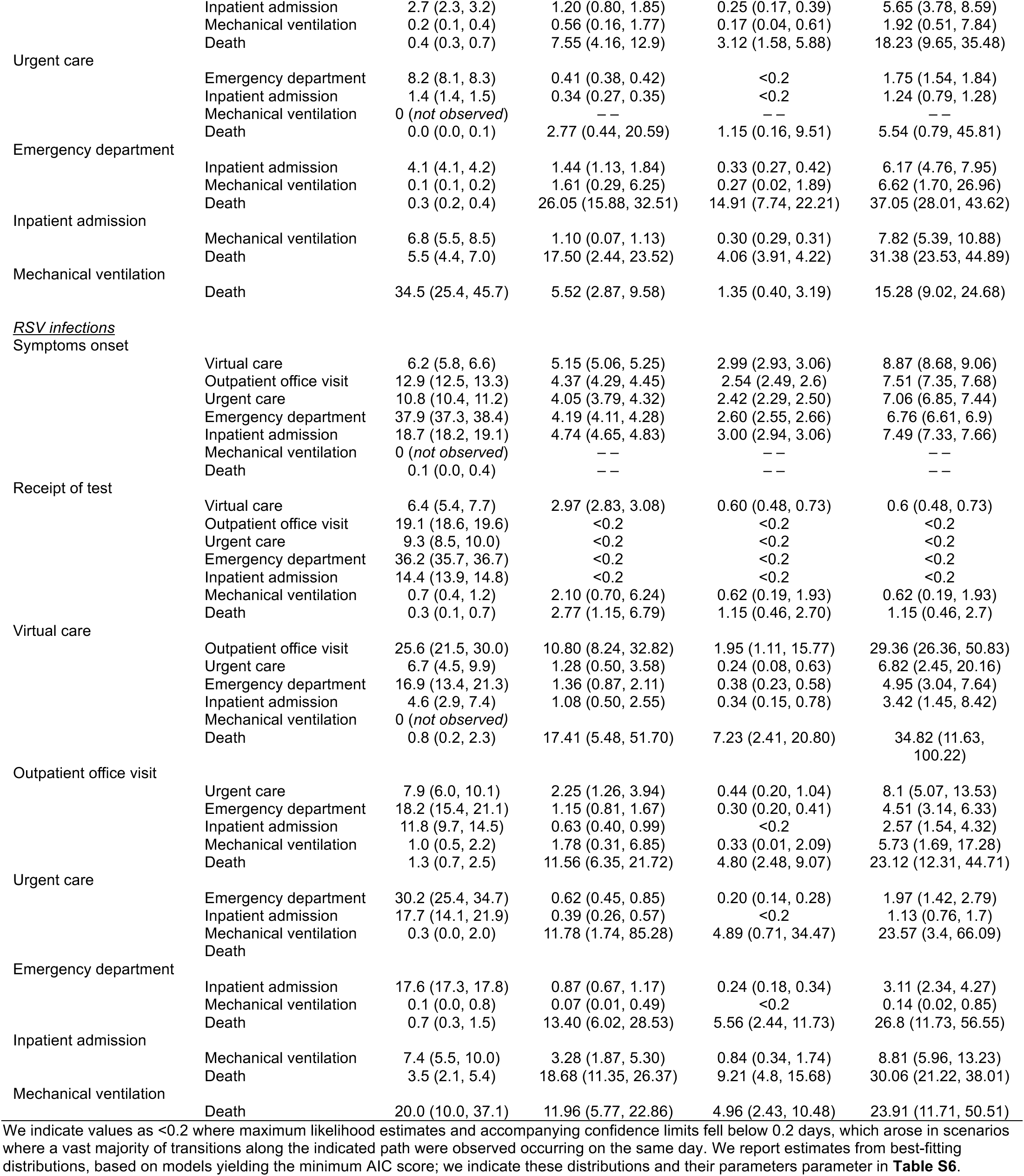
Care utilization pathways associated with each infecting virus using a follow-up period of 60 days.

**Table S6:**
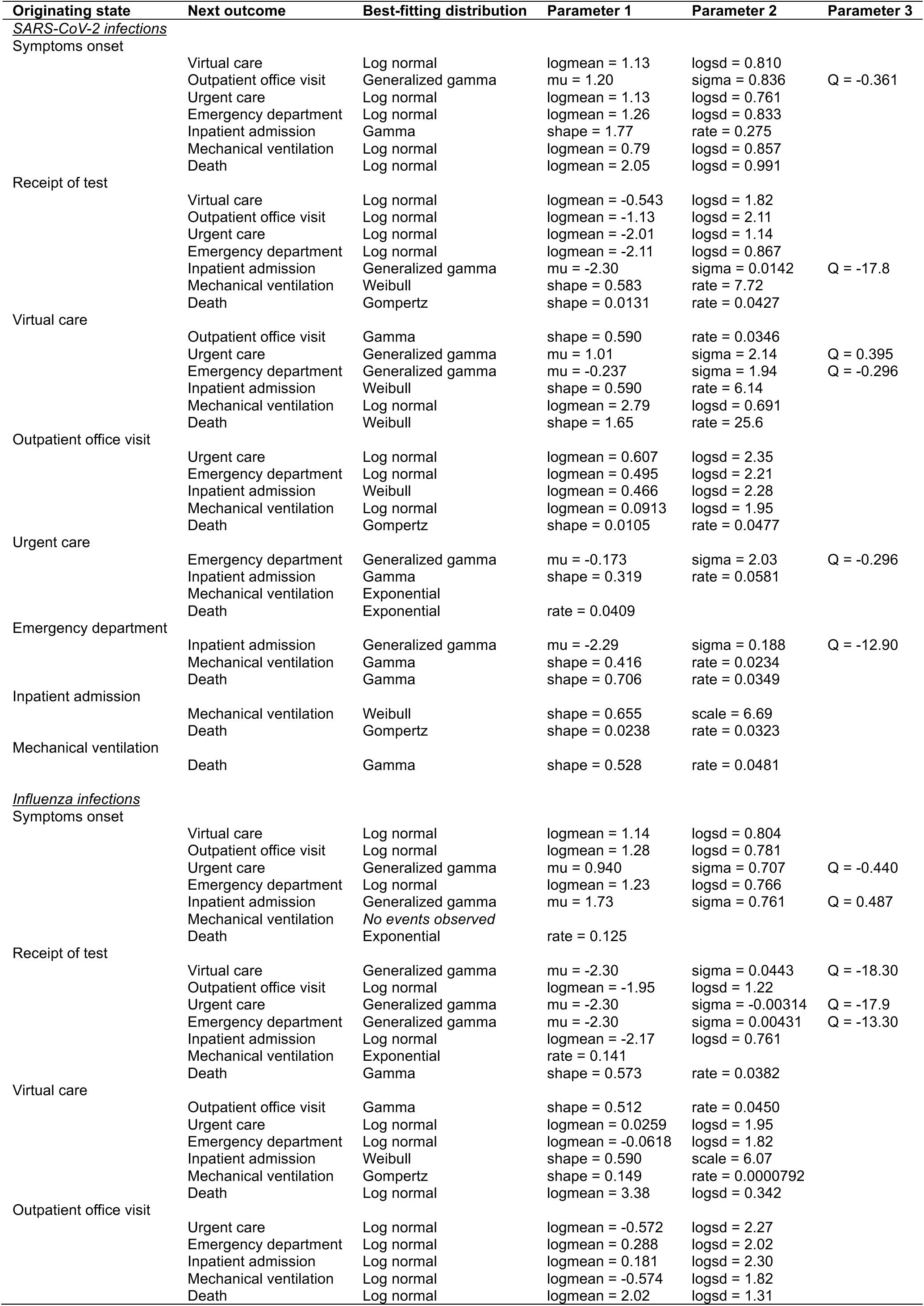
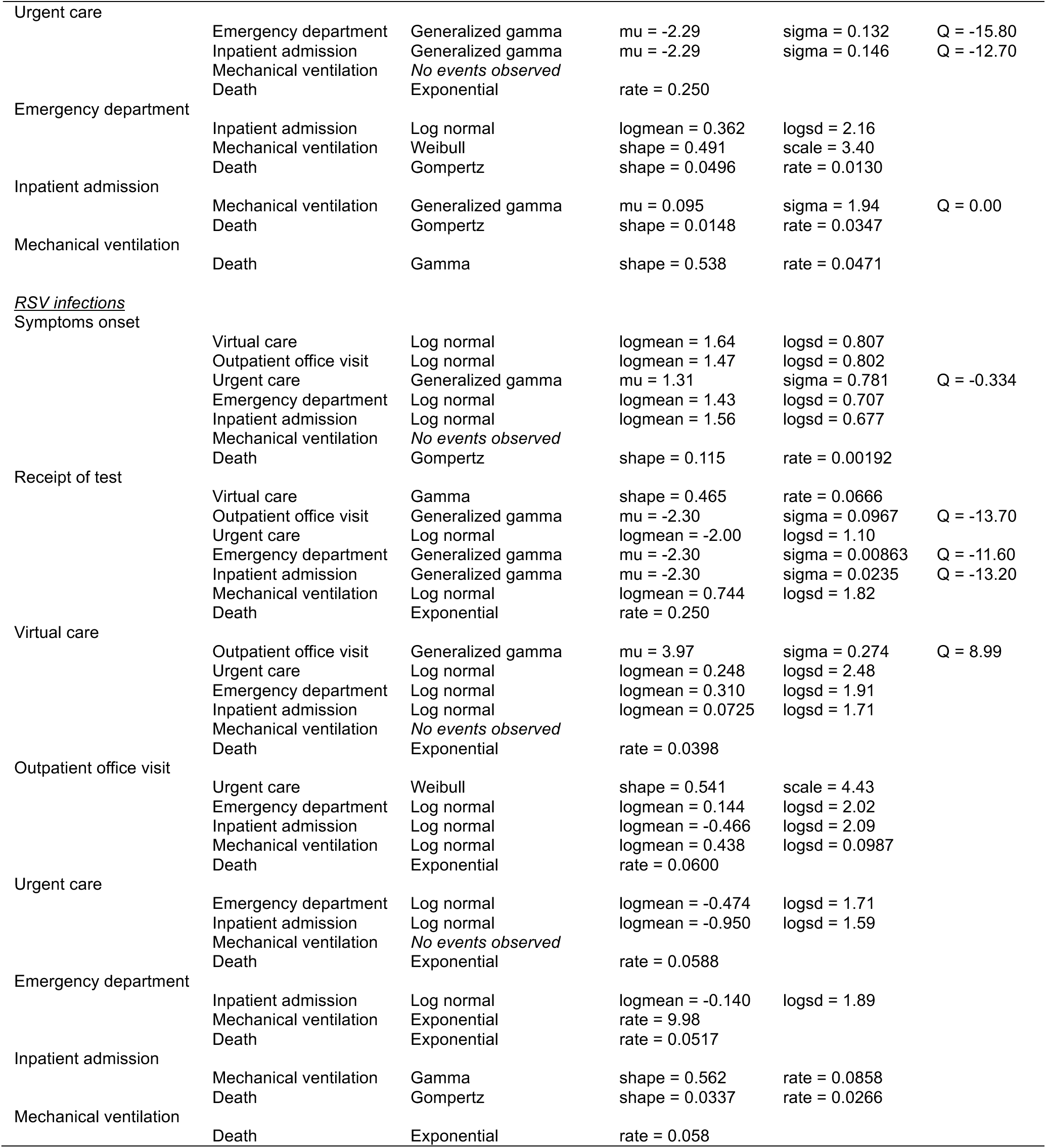
Best-fitting distributions for care utilization pathways for all infections using a 60 day follow-up period.

**Table S7:** Hospital length of stay estimates for admissions leading to discharge or mortality.

| Infection | Outcome | Probability, %<br>(95% CI) | Time to progression along indicated transition pathway, days (95% CI) |  |  |
| --- | --- | --- | --- | --- | --- |
|  |  |  | Median | 25%ile | 75%ile |
| <u>SARS-CoV-2 infections</u> |  |  |  |  |  |
|  | Any outcome | -- | 4.24 (4.14, 4.33) | 2.59 (2.53, 2.64) | 7.27 (7.09, 7.46) |
|  | Discharged alive | 93.1 (92.4, 93.7) | 4.11 (4.03, 4.20) | 2.55 (2.50, 2.61) | 6.94 (6.78, 7.12) |
|  | In-hospital mortality | 6.9 (6.3, 7.7) | 7.25 (6.47, 8.10) | 3.65 (3.21, 4.17) | 13.3 (12.05, 14.68) |
| <u>Influenza infections</u> |  |  |  |  |  |
|  | Any outcome | -- | 3.95 (3.78, 4.13) | 2.30 (2.19, 2.42) | 6.79 (6.47, 7.15) |
|  | Discharged alive | 96.0 (94.8, 96.8) | 3.91 (3.73, 4.09) | 2.30 (2.18, 2.42) | 6.65 (6.32, 6.99) |
|  | In-hospital mortality | 4.0 (3.2, 5.2) | 5.24 (3.93, 7.03) | 2.60 (1.83, 3.56) | 10.5 (7.74, 14.75) |
| <u>RSV infections</u> |  |  |  |  |  |
|  | Any outcome | -- | 4.33 (4.06, 4.61) | 2.54 (2.37, 2.72) | 7.39 (6.89, 7.91) |
|  | Discharged alive | 98.4 (96.9, 99.0) | 4.28 (4.01, 4.55) | 2.52 (2.35, 2.70) | 7.26 (6.77, 7.76) |
|  | In-hospital mortality | 1.6 (1.0, 3.1) | 11.3 (5.41, 16.51) | 5.80 (2.20, 10.46) | 17.6 (10.16, 23.71) |

**Table S8:**
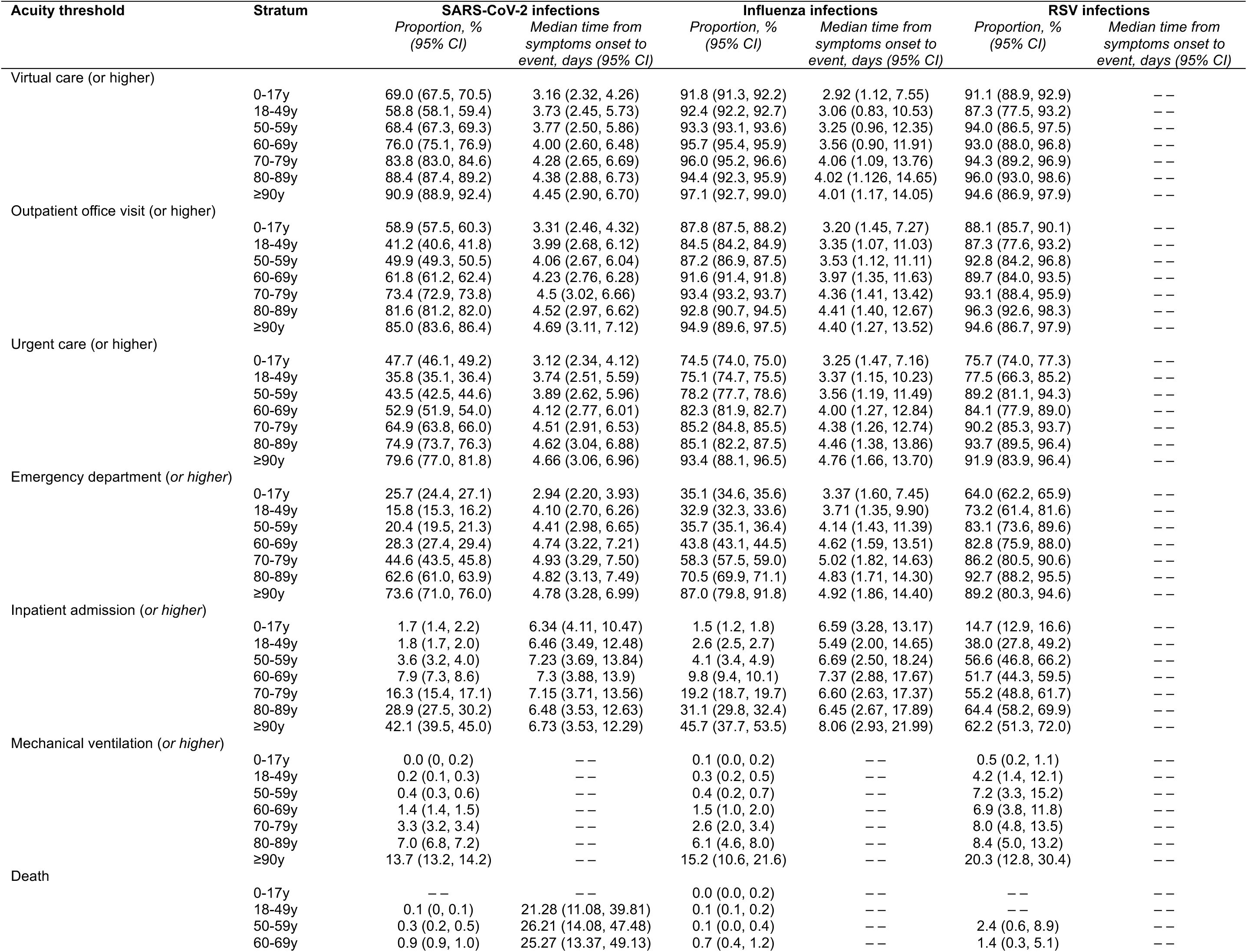
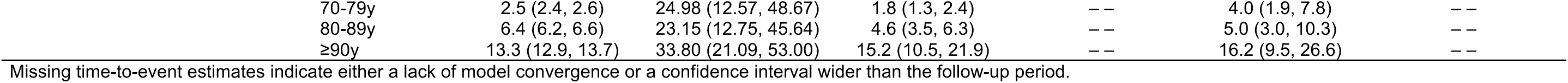
Proportions of cases attaining or exceeding each acuity threshold, by age.

| Acuity threshold | Stratum | SARS-CoV-2 infections |  | Influenza infections |  | RSV infections |  |
| --- | --- | --- | --- | --- | --- | --- | --- |
|  |  | Proportion, %<br>(95% CI) | Median time from<br>symptoms onset to<br>event, days (95% CI) | Proportion, %<br>(95% CI) | Median time from<br>symptoms onset to<br>event, days (95% CI) | Proportion, %<br>(95% CI) | Median time from<br>symptoms onset to<br>event, days (95% CI) |
| Virtual care (or higher) | 0-17y | 69.0 (67.5, 70.5) | 3.16 (2.32, 4.26) | 91.8 (91.3, 92.2) | 2.92 (1.12, 7.55) | 91.1 (88.9, 92.9) | -- |
|  | 18-49y | 58.8 (58.1, 59.4) | 3.73 (2.45, 5.73) | 92.4 (92.2, 92.7) | 3.06 (0.83, 10.53) | 87.3 (77.5, 93.2) | -- |
|  | 50-59y | 68.4 (67.3, 69.3) | 3.77 (2.50, 5.86) | 93.3 (93.1, 93.6) | 3.25 (0.96, 12.35) | 94.0 (86.5, 97.5) | -- |
|  | 60-69y | 76.0 (75.1, 76.9) | 4.00 (2.60, 6.48) | 95.7 (95.4, 95.9) | 3.56 (0.90, 11.91) | 93.0 (88.0, 96.8) | -- |
|  | 70-79y | 83.8 (83.0, 84.6) | 4.28 (2.65, 6.69) | 96.0 (95.2, 96.6) | 4.06 (1.09, 13.76) | 94.3 (89.2, 96.9) | -- |
|  | 80-89y | 88.4 (87.4, 89.2) | 4.38 (2.88, 6.73) | 94.4 (92.3, 95.9) | 4.02 (1.126, 14.65) | 96.0 (93.0, 98.6) | -- |
|  | ≥90y | 90.9 (88.9, 92.4) | 4.45 (2.90, 6.70) | 97.1 (92.7, 99.0) | 4.01 (1.17, 14.05) | 94.6 (86.9, 97.9) | -- |
| Outpatient office visit (or higher) | 0-17y | 58.9 (57.5, 60.3) | 3.31 (2.46, 4.32) | 87.8 (87.5, 88.2) | 3.20 (1.45, 7.27) | 88.1 (85.7, 90.1) | -- |
|  | 18-49y | 41.2 (40.6, 41.8) | 3.99 (2.68, 6.12) | 84.5 (84.2, 84.9) | 3.35 (1.07, 11.03) | 87.3 (77.6, 93.2) | -- |
|  | 50-59y | 49.9 (49.3, 50.5) | 4.06 (2.67, 6.04) | 87.2 (86.9, 87.5) | 3.53 (1.12, 11.11) | 92.8 (84.2, 96.8) | -- |
|  | 60-69y | 61.8 (61.2, 62.4) | 4.23 (2.76, 6.28) | 91.6 (91.4, 91.8) | 3.97 (1.35, 11.63) | 89.7 (84.0, 93.5) | -- |
|  | 70-79y | 73.4 (72.9, 73.8) | 4.5 (3.02, 6.66) | 93.4 (93.2, 93.7) | 4.36 (1.41, 13.42) | 93.1 (88.4, 95.9) | -- |
|  | 80-89y | 81.6 (81.2, 82.0) | 4.52 (2.97, 6.62) | 92.8 (90.7, 94.5) | 4.41 (1.40, 12.67) | 96.3 (92.6, 98.3) | -- |
|  | ≥90y | 85.0 (83.6, 86.4) | 4.69 (3.11, 7.12) | 94.9 (89.6, 97.5) | 4.40 (1.27, 13.52) | 94.6 (86.7, 97.9) | -- |
| Urgent care (or higher) | 0-17y | 47.7 (46.1, 49.2) | 3.12 (2.34, 4.12) | 74.5 (74.0, 75.0) | 3.25 (1.47, 7.16) | 75.7 (74.0, 77.3) | -- |
|  | 18-49y | 35.8 (35.1, 36.4) | 3.74 (2.51, 5.59) | 75.1 (74.7, 75.5) | 3.37 (1.15, 10.23) | 77.5 (66.3, 85.2) | -- |
|  | 50-59y | 43.5 (42.5, 44.6) | 3.89 (2.62, 5.96) | 78.2 (77.7, 78.6) | 3.56 (1.19, 11.49) | 89.2 (81.1, 94.3) | -- |
|  | 60-69y | 52.9 (51.9, 54.0) | 4.12 (2.77, 6.01) | 82.3 (81.9, 82.7) | 4.00 (1.27, 12.84) | 84.1 (77.9, 89.0) | -- |
|  | 70-79y | 64.9 (63.8, 66.0) | 4.51 (2.91, 6.53) | 85.2 (84.8, 85.5) | 4.38 (1.26, 12.74) | 90.2 (85.3, 93.7) | -- |
|  | 80-89y | 74.9 (73.7, 76.3) | 4.62 (3.04, 6.88) | 85.1 (82.2, 87.5) | 4.46 (1.38, 13.86) | 93.7 (89.5, 96.4) | -- |
|  | ≥90y | 79.6 (77.0, 81.8) | 4.66 (3.06, 6.96) | 93.4 (88.1, 96.5) | 4.76 (1.66, 13.70) | 91.9 (83.9, 96.4) | -- |
| Emergency department (or higher) | 0-17y | 25.7 (24.4, 27.1) | 2.94 (2.20, 3.93) | 35.1 (34.6, 35.6) | 3.37 (1.60, 7.45) | 64.0 (62.2, 65.9) | -- |
|  | 18-49y | 15.8 (15.3, 16.2) | 4.10 (2.70, 6.26) | 32.9 (32.3, 33.6) | 3.71 (1.35, 9.90) | 73.2 (61.4, 81.6) | -- |
|  | 50-59y | 20.4 (19.5, 21.3) | 4.41 (2.98, 6.65) | 35.7 (35.1, 36.4) | 4.14 (1.43, 11.39) | 83.1 (73.6, 89.6) | -- |
|  | 60-69y | 28.3 (27.4, 29.4) | 4.74 (3.22, 7.21) | 43.8 (43.1, 44.5) | 4.62 (1.59, 13.51) | 82.8 (75.9, 88.0) | -- |
|  | 70-79y | 44.6 (43.5, 45.8) | 4.93 (3.29, 7.50) | 58.3 (57.5, 59.0) | 5.02 (1.82, 14.63) | 86.2 (80.5, 90.6) | -- |
|  | 80-89y | 62.6 (61.0, 63.9) | 4.82 (3.13, 7.49) | 70.5 (69.9, 71.1) | 4.83 (1.71, 14.30) | 92.7 (88.2, 95.5) | -- |
|  | ≥90y | 73.6 (71.0, 76.0) | 4.78 (3.28, 6.99) | 87.0 (79.8, 91.8) | 4.92 (1.86, 14.40) | 89.2 (80.3, 94.6) | -- |
| Inpatient admission (or higher) | 0-17y | 1.7 (1.4, 2.2) | 6.34 (4.11, 10.47) | 1.5 (1.2, 1.8) | 6.59 (3.28, 13.17) | 14.7 (12.9, 16.6) | -- |
|  | 18-49y | 1.8 (1.7, 2.0) | 6.46 (3.49, 12.48) | 2.6 (2.5, 2.7) | 5.49 (2.00, 14.65) | 38.0 (27.8, 49.2) | -- |
|  | 50-59y | 3.6 (3.2, 4.0) | 7.23 (3.69, 13.84) | 4.1 (3.4, 4.9) | 6.69 (2.50, 18.24) | 56.6 (46.8, 66.2) | -- |
|  | 60-69y | 7.9 (7.3, 8.6) | 7.3 (3.88, 13.9) | 9.8 (9.4, 10.1) | 7.37 (2.88, 17.67) | 51.7 (44.3, 59.5) | -- |
|  | 70-79y | 16.3 (15.4, 17.1) | 7.15 (3.71, 13.56) | 19.2 (18.7, 19.7) | 6.60 (2.63, 17.37) | 55.2 (48.8, 61.7) | -- |
|  | 80-89y | 28.9 (27.5, 30.2) | 6.48 (3.53, 12.63) | 31.1 (29.8, 32.4) | 6.45 (2.67, 17.89) | 64.4 (58.2, 69.9) | -- |
|  | ≥90y | 42.1 (39.5, 45.0) | 6.73 (3.53, 12.29) | 45.7 (37.7, 53.5) | 8.06 (2.93, 21.99) | 62.2 (51.3, 72.0) | -- |
| Mechanical ventilation (or higher) | 0-17y | 0.0 (0, 0.2) | -- | 0.1 (0.0, 0.2) | -- | 0.5 (0.2, 1.1) | -- |
|  | 18-49y | 0.2 (0.1, 0.3) | -- | 0.3 (0.2, 0.5) | -- | 4.2 (1.4, 12.1) | -- |
|  | 50-59y | 0.4 (0.3, 0.6) | -- | 0.4 (0.2, 0.7) | -- | 7.2 (3.3, 15.2) | -- |
|  | 60-69y | 1.4 (1.4, 1.5) | -- | 1.5 (1.0, 2.0) | -- | 6.9 (3.8, 11.8) | -- |
|  | 70-79y | 3.3 (3.2, 3.4) | -- | 2.6 (2.0, 3.4) | -- | 8.0 (4.8, 13.5) | -- |
|  | 80-89y | 7.0 (6.8, 7.2) | -- | 6.1 (4.6, 8.0) | -- | 8.4 (5.0, 13.2) | -- |
|  | ≥90y | 13.7 (13.2, 14.2) | -- | 15.2 (10.6, 21.6) | -- | 20.3 (12.8, 30.4) | -- |
| Death | 0-17y | -- | -- | 0.0 (0.0, 0.2) | -- | -- | -- |
|  | 18-49y | 0.1 (0, 0.1) | 21.28 (11.08, 39.81) | 0.1 (0.1, 0.2) | -- | -- | -- |
|  | 50-59y | 0.3 (0.2, 0.5) | 26.21 (14.08, 47.48) | 0.1 (0.0, 0.4) | -- | 2.4 (0.6, 8.9) | -- |
|  | 60-69y | 0.9 (0.9, 1.0) | 25.27 (13.37, 49.13) | 0.7 (0.4, 1.2) | -- | 1.4 (0.3, 5.1) | -- |

|  |  |  |  |  |  |  |
| --- | --- | --- | --- | --- | --- | --- |
| 70-79y | 2.5 (2.4, 2.6) | 24.98 (12.57, 48.67) | 1.8 (1.3, 2.4) | -- | 4.0 (1.9, 7.8) | -- |
| 80-89y | 6.4 (6.2, 6.6) | 23.15 (12.75, 45.64) | 4.6 (3.5, 6.3) | -- | 5.0 (3.0, 10.3) | -- |
| ≥90y | 13.3 (12.9, 13.7) | 33.80 (21.09, 53.00) | 15.2 (10.5, 21.9) | -- | 16.2 (9.5, 26.6) | -- |

Missing time-to-event estimates indicate either a lack of model convergence or a confidence interval wider than the follow-up period.

**Table S9:**
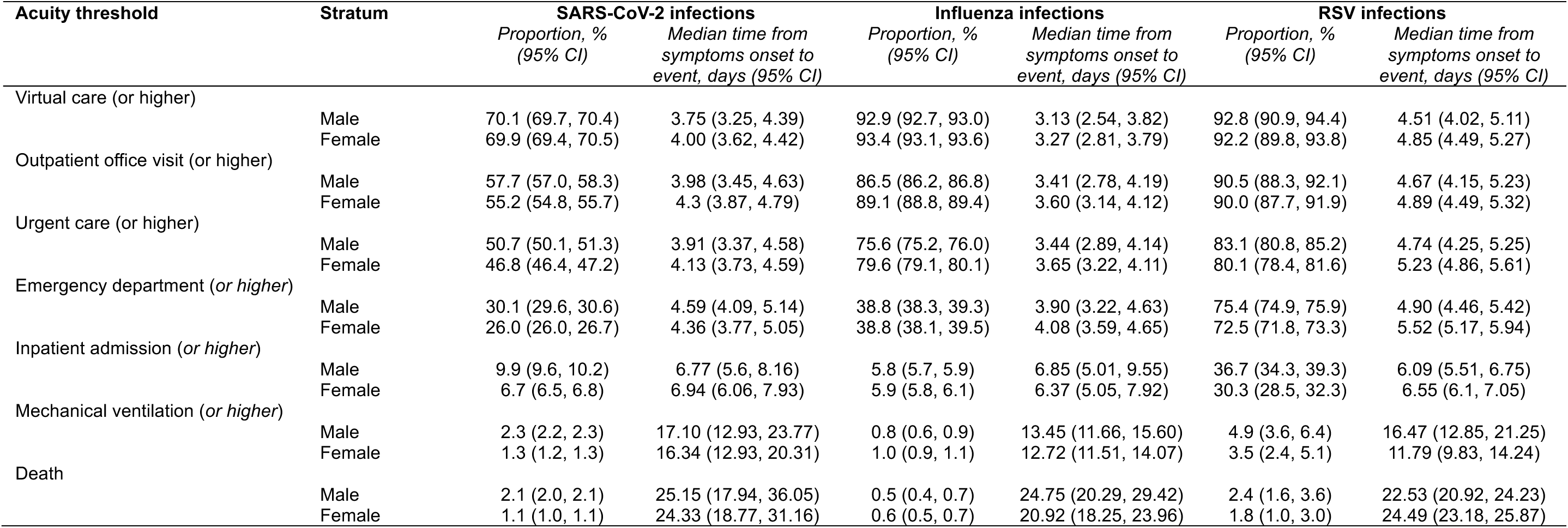
Proportions of cases attaining or exceeding each acuity threshold, by sex.

| Acuity threshold | Stratum | SARS-CoV-2 infections |  | Influenza infections |  | RSV infections |  |
| --- | --- | --- | --- | --- | --- | --- | --- |
|  |  | Proportion, %<br>(95% CI) | Median time from<br>symptoms onset to<br>event, days (95% CI) | Proportion, %<br>(95% CI) | Median time from<br>symptoms onset to<br>event, days (95% CI) | Proportion, %<br>(95% CI) | Median time from<br>symptoms onset to<br>event, days (95% CI) |
| Virtual care (or higher) | Male | 70.1 (69.7, 70.4) | 3.75 (3.25, 4.39) | 92.9 (92.7, 93.0) | 3.13 (2.54, 3.82) | 92.8 (90.9, 94.4) | 4.51 (4.02, 5.11) |
|  | Female | 69.9 (69.4, 70.5) | 4.00 (3.62, 4.42) | 93.4 (93.1, 93.6) | 3.27 (2.81, 3.79) | 92.2 (89.8, 93.8) | 4.85 (4.49, 5.27) |
| Outpatient office visit (or higher) | Male | 57.7 (57.0, 58.3) | 3.98 (3.45, 4.63) | 86.5 (86.2, 86.8) | 3.41 (2.78, 4.19) | 90.5 (88.3, 92.1) | 4.67 (4.15, 5.23) |
|  | Female | 55.2 (54.8, 55.7) | 4.3 (3.87, 4.79) | 89.1 (88.8, 89.4) | 3.60 (3.14, 4.12) | 90.0 (87.7, 91.9) | 4.89 (4.49, 5.32) |
| Urgent care (or higher) | Male | 50.7 (50.1, 51.3) | 3.91 (3.37, 4.58) | 75.6 (75.2, 76.0) | 3.44 (2.89, 4.14) | 83.1 (80.8, 85.2) | 4.74 (4.25, 5.25) |
|  | Female | 46.8 (46.4, 47.2) | 4.13 (3.73, 4.59) | 79.6 (79.1, 80.1) | 3.65 (3.22, 4.11) | 80.1 (78.4, 81.6) | 5.23 (4.86, 5.61) |
| Emergency department (or higher) | Male | 30.1 (29.6, 30.6) | 4.59 (4.09, 5.14) | 38.8 (38.3, 39.3) | 3.90 (3.22, 4.63) | 75.4 (74.9, 75.9) | 4.90 (4.46, 5.42) |
|  | Female | 26.0 (26.0, 26.7) | 4.36 (3.77, 5.05) | 38.8 (38.1, 39.5) | 4.08 (3.59, 4.65) | 72.5 (71.8, 73.3) | 5.52 (5.17, 5.94) |
| Inpatient admission (or higher) | Male | 9.9 (9.6, 10.2) | 6.77 (5.6, 8.16) | 5.8 (5.7, 5.9) | 6.85 (5.01, 9.55) | 36.7 (34.3, 39.3) | 6.09 (5.51, 6.75) |
|  | Female | 6.7 (6.5, 6.8) | 6.94 (6.06, 7.93) | 5.9 (5.8, 6.1) | 6.37 (5.05, 7.92) | 30.3 (28.5, 32.3) | 6.55 (6.1, 7.05) |
| Mechanical ventilation (or higher) | Male | 2.3 (2.2, 2.3) | 17.10 (12.93, 23.77) | 0.8 (0.6, 0.9) | 13.45 (11.66, 15.60) | 4.9 (3.6, 6.4) | 16.47 (12.85, 21.25) |
|  | Female | 1.3 (1.2, 1.3) | 16.34 (12.93, 20.31) | 1.0 (0.9, 1.1) | 12.72 (11.51, 14.07) | 3.5 (2.4, 5.1) | 11.79 (9.83, 14.24) |
| Death | Male | 2.1 (2.0, 2.1) | 25.15 (17.94, 36.05) | 0.5 (0.4, 0.7) | 24.75 (20.29, 29.42) | 2.4 (1.6, 3.6) | 22.53 (20.92, 24.23) |
|  | Female | 1.1 (1.0, 1.1) | 24.33 (18.77, 31.16) | 0.6 (0.5, 0.7) | 20.92 (18.25, 23.96) | 1.8 (1.0, 3.0) | 24.49 (23.18, 25.87) |

**Table S10:** Proportions of cases attaining or exceeding each acuity threshold, by neighborhood deprivation index.

| Acuity threshold | Stratum | SARS-CoV-2 infections |  | Influenza infections |  | RSV infections |  |
| --- | --- | --- | --- | --- | --- | --- | --- |
|  |  | Proportion, %<br>(95% CI) | Median time from<br>symptoms onset to<br>event, days (95% CI) | Proportion, %<br>(95% CI) | Median time from<br>symptoms onset to<br>event, days (95% CI) | Proportion, %<br>(95% CI) | Median time from<br>symptoms onset to<br>event, days (95% CI) |
| Virtual care (or higher) | Below -1 | 70.0 (69.5, 70.5) | 4.02 (3.78, 4.28) | 91.0 (89.0, 92.7) | 3.39 (3.09, 3.71) | 88.2 (79.9, 93.6) | 5.11 (3.86, 6.73) |
|  | -1 ≤ NDI < 0 | 71.3 (69.5, 73.0) | 4.00 (3.66, 4.38) | 92.4 (92.1, 92.7) | 3.27 (2.89, 3.70) | 94.0 (91.5, 95.6) | 4.92 (3.24, 7.40) |
|  | 0 ≤ NDI < 1 | 70.1 (69.6, 70.6) | 3.84 (3.53, 4.17) | 93.7 (93.5, 93.9) | 3.21 (2.83, 3.65) | 93.7 (91.4, 95.4) | 4.33 (3.01, 6.32) |
|  | Above 1 | 69.5 (68.9, 70.0) | 3.78 (3.46, 4.13) | 93.3 (93.0, 93.6) | 3.09 (2.73, 3.53) | 91.4 (87.1, 94.2) | 4.85 (3.30, 7.01) |
| Outpatient office visit (or higher) | Below -1 | 55.6 (54.9, 56.2) | 4.25 (4, 4.51) | 84.9 (82.4, 87.2) | 3.82 (3.51, 4.17) | 82.9 (73.4, 89.7) | 5.21 (4.06, 6.68) |
|  | -1 ≤ NDI < 0 | 59.0 (58.6, 59.4) | 4.32 (3.93, 4.7) | 86.9 (86.5, 87.3) | 3.56 (3.15, 4.03) | 91.4 (89.3, 93.3) | 5.00 (3.50, 7.18) |
|  | 0 ≤ NDI < 1 | 56.6 (56.1, 57.2) | 4.1 (3.72, 4.49) | 88.2 (87.8, 88.6) | 3.53 (3.14, 3.96) | 92.3 (90.2, 93.9) | 4.48 (3.07, 6.53) |
|  | Above 1 | 55.3 (54.7, 55.9) | 3.99 (3.64, 4.36) | 88.6 (88.2, 89.0) | 3.33 (2.96, 3.77) | 89.1 (85.0, 92.1) | 4.89 (3.39, 7.11) |
| Urgent care (or higher) | Below -1 | 47.4 (46.8, 48.0) | 4.14 (3.89, 4.38) | 72.0 (68.8, 74.7) | 3.88 (3.61, 4.17) | 75.0 (63.7, 83.4) | 5.22 (4.17, 6.73) |
|  | -1 ≤ NDI < 0 | 49.9 (49.4, 50.3) | 4.18 (3.82, 4.57) | 75.6 (75.0, 76.2) | 3.59 (3.22, 4.00) | 82.3 (79.6, 84.7) | 5.13 (3.76, 7.14) |
|  | 0 ≤ NDI < 1 | 47.8 (47.2, 48.5) | 4.01 (3.67, 4.39) | 78.3 (77.7, 78.8) | 3.55 (3.20, 3.96) | 83.6 (81.1, 85.8) | 4.68 (3.44, 6.59) |
|  | Above 1 | 47.5 (46.8, 48.1) | 3.82 (3.48, 4.16) | 79.3 (78.8, 79.9) | 3.43 (3.08, 3.81) | 82.0 (77.6, 85.8) | 5.12 (3.68, 7.36) |
| Emergency department (or higher) | Below -1 | 27.5 (27.0, 28.1) | 4.55 (4.26, 4.88) | 35.4 (32.2, 38.8) | 4.35 (4.00, 4.69) | 69.7 (58.0, 78.5) | 5.64 (4.64, 6.98) |
|  | -1 ≤ NDI < 0 | 33.2 (32.8, 33.6) | 4.71 (4.34, 5.22) | 36.4 (35.6, 37.1) | 3.98 (3.55, 4.42) | 73.7 (70.5, 76.5) | 5.42 (3.96, 7.34) |
|  | 0 ≤ NDI < 1 | 28.3 (27.8, 28.8) | 4.47 (4.06, 4.91) | 39.6 (38.8, 40.3) | 4.12 (3.70, 4.59) | 75.9 (73.2, 78.5) | 4.88 (3.60, 6.71) |
|  | Above 1 | 26.5 (26.0, 27.0) | 4.28 (3.90, 4.70) | 39.7 (38.9, 40.5) | 3.89 (3.49, 4.33) | 74.5 (69.0, 79.1) | 5.28 (3.86, 7.22) |
| Inpatient admission (or higher) | Below -1 | 8.5 (8.2, 8.7) | 6.92 (6.39, 7.5) | 5.3 (4.0, 6.9) | 7.59 (6.69, 8.64) | 35.5 (25.5, 47.4) | 6.46 (5.15, 8.16) |
|  | -1 ≤ NDI < 0 | 9.7 (8.6, 11.0) | 6.68 (5.94, 7.44) | 6.1 (5.9, 6.2) | 6.29 (5.20, 7.72) | 31.2 (28.2, 34.4) | 6.75 (4.91, 9.29) |
|  | 0 ≤ NDI < 1 | 8.8 (8.5, 9.0) | 6.96 (6.24, 7.79) | 6.2 (6.0, 6.3) | 6.76 (5.64, 8.16) | 36.0 (33.2, 38.9) | 6.01 (4.39, 8.23) |
|  | Above 1 | 7.7 (7.4, 7.9) | 7.02 (6.29, 7.88) | 5.4 (5.2, 5.5) | 6.77 (5.64, 8.08) | 34.1 (29.1, 39.4) | 5.94 (4.31, 8.19) |
| Mechanical ventilation (or higher) | Below -1 | 1.8 (1.7, 1.8) | 14.63 (12.59, 16.92) | 0.5 (0.2, 1.3) | 21.95 (16.19, 29.16) | 3.9 (1.2, 12.2) | 17.86 (10.28, 31.21) |
|  | -1 ≤ NDI < 0 | 2.2 (1.7, 2.8) | 17.41 (14.05, 21.27) | 0.8 (0.6, 1.0) | 13.30 (8.59, 20.35) | 4.1 (2.8, 6.3) | 14.86 (7.27, 30.00) |
|  | 0 ≤ NDI < 1 | 1.7 (1.6, 1.7) | 16.18 (13.13, 20.04) | 1.0 (0.8, 1.2) | 11.91 (7.66, 18.37) | 4.1 (2.7, 6.2) | 13.84 (6.72, 29.12) |
|  | Above 1 | 1.7 (1.6, 1.7) | 16.96 (13.81, 21.01) | 0.9 (0.7, 1.2) | 14.54 (9.18, 22.65) | 4.5 (2.5, 7.7) | 10.20 (4.81, 21.43) |
| Death | Below -1 | 1.5 (1.5, 1.6) | 20.73 (17.49, 24.50) | 0.3 (0.1, 1.1) | 23.63 (16.16, 34.79) | 2.6 (0.7, 10.7) | 25.20 (11.69, 50.61) |
|  | -1 ≤ NDI < 0 | 2.0 (1.5, 2.8) | 25.92 (20.50, 32.27) | 0.6 (0.5, 0.8) | 23.03 (13.52, 38.35) | 2.7 (1.6, 4.6) | 20.13 (7.10, 52.93) |
|  | 0 ≤ NDI < 1 | 1.5 (1.5, 1.6) | 24.01 (19.30, 30.12) | 0.6 (0.4, 0.7) | 22.15 (12.90, 38.04) | 2.0 (1.1, 3.5) | 27.01 (9.31, 73.35) |
|  | Above 1 | 1.5 (1.4, 1.5) | 26.86 (21.39, 33.29) | 0.5 (0.3, 0.8) | 23.18 (13.72, 39.69) | 1.1 (0.4, 3.3) | 28.15 (10.09, 77.42) |

**Table S11:**
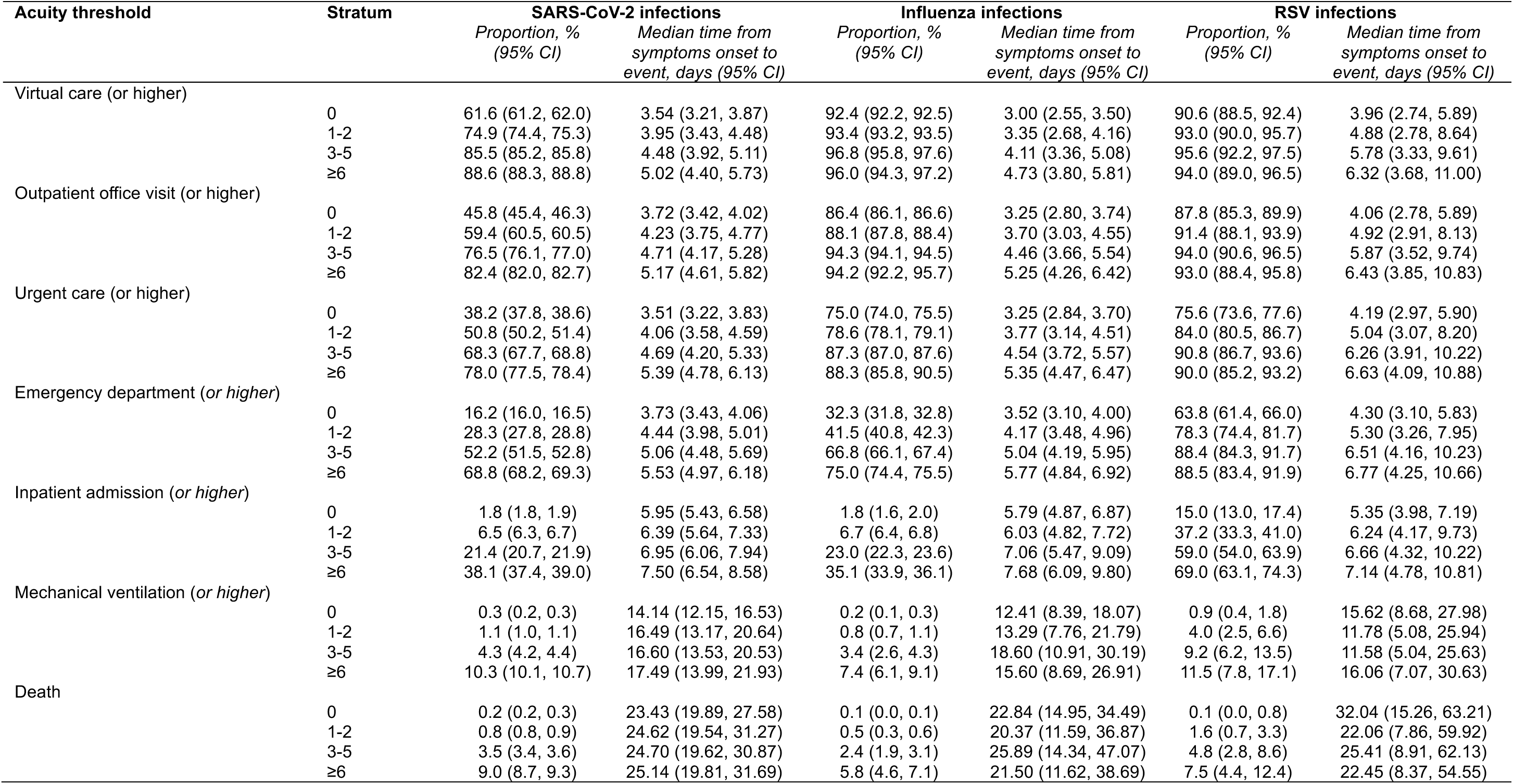
Proportions of cases attaining or exceeding each acuity threshold, by Charlson comorbidty index values.

| Acuity threshold | Stratum | SARS-CoV-2 infections |  | Influenza infections |  | RSV infections |  |
| --- | --- | --- | --- | --- | --- | --- | --- |
|  |  | Proportion, %<br>(95% CI) | Median time from<br>symptoms onset to<br>event, days (95% CI) | Proportion, %<br>(95% CI) | Median time from<br>symptoms onset to<br>event, days (95% CI) | Proportion, %<br>(95% CI) | Median time from<br>symptoms onset to<br>event, days (95% CI) |
| Virtual care (or higher) | 0 | 61.6 (61.2, 62.0) | 3.54 (3.21, 3.87) | 92.4 (92.2, 92.5) | 3.00 (2.55, 3.50) | 90.6 (88.5, 92.4) | 3.96 (2.74, 5.89) |
|  | 1-2 | 74.9 (74.4, 75.3) | 3.95 (3.43, 4.48) | 93.4 (93.2, 93.5) | 3.35 (2.68, 4.16) | 93.0 (90.0, 95.7) | 4.88 (2.78, 8.64) |
|  | 3-5 | 85.5 (85.2, 85.8) | 4.48 (3.92, 5.11) | 96.8 (95.8, 97.6) | 4.11 (3.36, 5.08) | 95.6 (92.2, 97.5) | 5.78 (3.33, 9.61) |
|  | ≥6 | 88.6 (88.3, 88.8) | 5.02 (4.40, 5.73) | 96.0 (94.3, 97.2) | 4.73 (3.80, 5.81) | 94.0 (89.0, 96.5) | 6.32 (3.68, 11.00) |
| Outpatient office visit (or higher) | 0 | 45.8 (45.4, 46.3) | 3.72 (3.42, 4.02) | 86.4 (86.1, 86.6) | 3.25 (2.80, 3.74) | 87.8 (85.3, 89.9) | 4.06 (2.78, 5.89) |
|  | 1-2 | 59.4 (60.5, 60.5) | 4.23 (3.75, 4.77) | 88.1 (87.8, 88.4) | 3.70 (3.03, 4.55) | 91.4 (88.1, 93.9) | 4.92 (2.91, 8.13) |
|  | 3-5 | 76.5 (76.1, 77.0) | 4.71 (4.17, 5.28) | 94.3 (94.1, 94.5) | 4.46 (3.66, 5.54) | 94.0 (90.6, 96.5) | 5.87 (3.52, 9.74) |
|  | ≥6 | 82.4 (82.0, 82.7) | 5.17 (4.61, 5.82) | 94.2 (92.2, 95.7) | 5.25 (4.26, 6.42) | 93.0 (88.4, 95.8) | 6.43 (3.85, 10.83) |
| Urgent care (or higher) | 0 | 38.2 (37.8, 38.6) | 3.51 (3.22, 3.83) | 75.0 (74.0, 75.5) | 3.25 (2.84, 3.70) | 75.6 (73.6, 77.6) | 4.19 (2.97, 5.90) |
|  | 1-2 | 50.8 (50.2, 51.4) | 4.06 (3.58, 4.59) | 78.6 (78.1, 79.1) | 3.77 (3.14, 4.51) | 84.0 (80.5, 86.7) | 5.04 (3.07, 8.20) |
|  | 3-5 | 68.3 (67.7, 68.8) | 4.69 (4.20, 5.33) | 87.3 (87.0, 87.6) | 4.54 (3.72, 5.57) | 90.8 (86.7, 93.6) | 6.26 (3.91, 10.22) |
|  | ≥6 | 78.0 (77.5, 78.4) | 5.39 (4.78, 6.13) | 88.3 (85.8, 90.5) | 5.35 (4.47, 6.47) | 90.0 (85.2, 93.2) | 6.63 (4.09, 10.88) |
| Emergency department (or higher) | 0 | 16.2 (16.0, 16.5) | 3.73 (3.43, 4.06) | 32.3 (31.8, 32.8) | 3.52 (3.10, 4.00) | 63.8 (61.4, 66.0) | 4.30 (3.10, 5.83) |
|  | 1-2 | 28.3 (27.8, 28.8) | 4.44 (3.98, 5.01) | 41.5 (40.8, 42.3) | 4.17 (3.48, 4.96) | 78.3 (74.4, 81.7) | 5.30 (3.26, 7.95) |
|  | 3-5 | 52.2 (51.5, 52.8) | 5.06 (4.48, 5.69) | 66.8 (66.1, 67.4) | 5.04 (4.19, 5.95) | 88.4 (84.3, 91.7) | 6.51 (4.16, 10.23) |
|  | ≥6 | 68.8 (68.2, 69.3) | 5.53 (4.97, 6.18) | 75.0 (74.4, 75.5) | 5.77 (4.84, 6.92) | 88.5 (83.4, 91.9) | 6.77 (4.25, 10.66) |
| Inpatient admission (or higher) | 0 | 1.8 (1.8, 1.9) | 5.95 (5.43, 6.58) | 1.8 (1.6, 2.0) | 5.79 (4.87, 6.87) | 15.0 (13.0, 17.4) | 5.35 (3.98, 7.19) |
|  | 1-2 | 6.5 (6.3, 6.7) | 6.39 (5.64, 7.33) | 6.7 (6.4, 6.8) | 6.03 (4.82, 7.72) | 37.2 (33.3, 41.0) | 6.24 (4.17, 9.73) |
|  | 3-5 | 21.4 (20.7, 21.9) | 6.95 (6.06, 7.94) | 23.0 (22.3, 23.6) | 7.06 (5.47, 9.09) | 59.0 (54.0, 63.9) | 6.66 (4.32, 10.22) |
|  | ≥6 | 38.1 (37.4, 39.0) | 7.50 (6.54, 8.58) | 35.1 (33.9, 36.1) | 7.68 (6.09, 9.80) | 69.0 (63.1, 74.3) | 7.14 (4.78, 10.81) |
| Mechanical ventilation (or higher) | 0 | 0.3 (0.2, 0.3) | 14.14 (12.15, 16.53) | 0.2 (0.1, 0.3) | 12.41 (8.39, 18.07) | 0.9 (0.4, 1.8) | 15.62 (8.68, 27.98) |
|  | 1-2 | 1.1 (1.0, 1.1) | 16.49 (13.17, 20.64) | 0.8 (0.7, 1.1) | 13.29 (7.76, 21.79) | 4.0 (2.5, 6.6) | 11.78 (5.08, 25.94) |
|  | 3-5 | 4.3 (4.2, 4.4) | 16.60 (13.53, 20.53) | 3.4 (2.6, 4.3) | 18.60 (10.91, 30.19) | 9.2 (6.2, 13.5) | 11.58 (5.04, 25.63) |
|  | ≥6 | 10.3 (10.1, 10.7) | 17.49 (13.99, 21.93) | 7.4 (6.1, 9.1) | 15.60 (8.69, 26.91) | 11.5 (7.8, 17.1) | 16.06 (7.07, 30.63) |
| Death | 0 | 0.2 (0.2, 0.3) | 23.43 (19.89, 27.58) | 0.1 (0.0, 0.1) | 22.84 (14.95, 34.49) | 0.1 (0.0, 0.8) | 32.04 (15.26, 63.21) |
|  | 1-2 | 0.8 (0.8, 0.9) | 24.62 (19.54, 31.27) | 0.5 (0.3, 0.6) | 20.37 (11.59, 36.87) | 1.6 (0.7, 3.3) | 22.06 (7.86, 59.92) |
|  | 3-5 | 3.5 (3.4, 3.6) | 24.70 (19.62, 30.87) | 2.4 (1.9, 3.1) | 25.89 (14.34, 47.07) | 4.8 (2.8, 8.6) | 25.41 (8.91, 62.13) |
|  | ≥6 | 9.0 (8.7, 9.3) | 25.14 (19.81, 31.69) | 5.8 (4.6, 7.1) | 21.50 (11.62, 38.69) | 7.5 (4.4, 12.4) | 22.45 (8.37, 54.55) |

**Table S12:** Proportions of cases attaining or exceeding each acuity threshold, by vaccination status.

| Acuity threshold | Stratum | SARS-CoV-2 infections |  | Influenza infections |  | RSV infections |  |
| --- | --- | --- | --- | --- | --- | --- | --- |
|  |  | Proportion, %<br>(95% CI) | Median time from<br>symptoms onset to<br>event, days (95% CI) | Proportion, %<br>(95% CI) | Median time from<br>symptoms onset to<br>event, days (95% CI) | Proportion, %<br>(95% CI) | Median time from<br>symptoms onset to<br>event, days (95% CI) |
| Virtual care (or higher) | Unvaccinated | 72.8 (72.3, 73.3) | 3.66 (3.22, 4.10) | 92.6 (92.4, 92.8) | 3.04 (2.60, 3.47) | 92.4 (91.1, 93.7) | 4.69 (3.39, 6.59) |
|  | Vaccinated<sup>1</sup> | 68.4 (68.0, 68.7) | 3.70 (3.39, 4.05) | 93.5 (93.2, 93.7) | 3.35 (2.73, 4.09) | 96.3 (76.5, 99.5) | 4.65 (2.88, 7.31) |
|  | COVID-19: ≥3 doses | 70.0 (69.5, 70.5) | 4.00 (3.53, 4.51) | -- | -- | -- | -- |
| Outpatient office visit (or higher) | Unvaccinated | 61.3 (60.7, 61.9) | 3.83 (3.42, 4.31) | 86.7 (86.5, 87.0) | 3.32 (2.90, 3.75) | 90.2 (88.6, 91.5) | 4.79 (3.46, 6.58) |
|  | Vaccinated<sup>1</sup> | 55.8 (55.4, 56.2) | 3.87 (3.55, 4.21) | 88.5 (88.1, 88.8) | 3.68 (3.02, 4.41) | 96.3 (79.0, 99.5) | 4.65 (2.90, 7.71) |
|  | COVID-19: ≥3 doses | 55.6 (54.9, 56.2) | 4.31 (3.86, 4.87) | -- | -- | -- | -- |
| Urgent care (or higher) | Unvaccinated | 53.6 (53.0, 54.2) | 3.73 (3.32, 4.23) | 75.9 (75.6, 76.4) | 3.35 (2.97, 3.73) | 81.6 (81.4, 81.8) | 5.00 (3.82, 6.35) |
|  | Vaccinated<sup>1</sup> | 48.5 (48.0, 48.9) | 3.77 (3.45, 4.14) | 78.6 (78.0, 79.0) | 3.72 (3.13, 4.42) | 88.9 (72.5, 96.2) | 5.13 (3.52, 7.20) |
|  | COVID-19: ≥3 doses | 47.4 (46.8, 48.0) | 4.17 (3.66, 4.77) | -- | -- | -- | -- |
| Emergency department (or higher) | Unvaccinated | 33.3 (32.7, 33.8) | 3.98 (3.53, 4.46) | 29.2 (28.7, 29.6) | 3.87 (3.47, 4.34) | 74.0 (73.5, 74.5) | 5.22 (4.21, 6.43) |
|  | Vaccinated<sup>1</sup> | 25.8 (25.4, 26.1) | 4.19 (3.85, 4.55) | 46.7 (46.0, 47.4) | 4.07 (3.46, 4.76) | 77.9 (59.9, 89.5) | 5.78 (4.33, 7.75) |
|  | COVID-19: ≥3 doses | 27.5 (27.0, 28.1) | 4.67 (4.16, 5.26) | -- | -- | -- | -- |
| Inpatient admission (or higher) | Unvaccinated | 7.7 (7.4, 7.9) | 6.44 (5.68, 7.31) | 3.1 (2.8, 3.5) | 6.64 (5.18, 8.77) | 33.7 (33.3, 34.0) | 6.35 (5.23, 7.67) |
|  | Vaccinated<sup>1</sup> | 6.2 (6.1, 6.4) | 6.1 (5.59, 6.71) | 8.1 (7.9, 8.3) | 6.44 (5.30, 7.85) | 40.7 (24.5, 59.3) | 6.90 (5.32, 9.11) |
|  | COVID-19: ≥3 doses | 8.5 (8.2, 8.7) | 7.07 (6.25, 8.04) | -- | -- | -- | -- |
| Mechanical ventilation (or higher) | Unvaccinated | 1.4 (1.3, 1.5) | 16.17 (13.81, 18.82) | 0.4 (0.3, 0.6) | 12.74 (11.63, 13.99) | 4.3 (3.3, 5.3) | 13.21 (8.61, 20.48) |
|  | Vaccinated<sup>1</sup> | 1.3 (1.2, 1.5) | 15.62 (12.49, 19.41) | 1.2 (1.1, 1.3) | 16.12 (14.07, 18.43) | 3.7 (0.5, 19.2) | 33.81 (18.68, 62.50) |
|  | COVID-19: ≥3 doses | 1.8 (1.7, 1.8) | 17.02 (13.58, 21.03) | -- | -- | -- | -- |
| Death | Unvaccinated | 1.0 (1.0, 1.9) | 23.01 (17.83, 29.09) | 0.3 (0.2, 0.4) | 20.29 (18.11, 22.71) | 2.2 (1.5, 3.0) | -- |
|  | Vaccinated<sup>1</sup> | 1.2 (1.0, 1.4) | 24.54 (20.71, 29.15) | 0.8 (0.7, 0.8) | 23.37 (19.96, 27.26) | -- | -- |
|  | COVID-19: ≥3 doses | 1.5 (1.5, 1.6) | 25.07 (19.45, 32.17) | -- | -- | -- | -- |

<sup>1</sup>For SARS-CoV-2 infections, rows correspond to a category of “under-vaccinated” individuals who received 1-2 total COVID-19 vaccine doses. For influenza infections, individuals are considered vaccinated if they received seasonal influenza vaccine for the applicable season; for RSV infections, individuals are considered vaccinated if they received any RSV vaccine. Missing time-to-event estimates indicate either a lack of model convergence or a confidence interval wider than the follow-up period

**Table S13:**
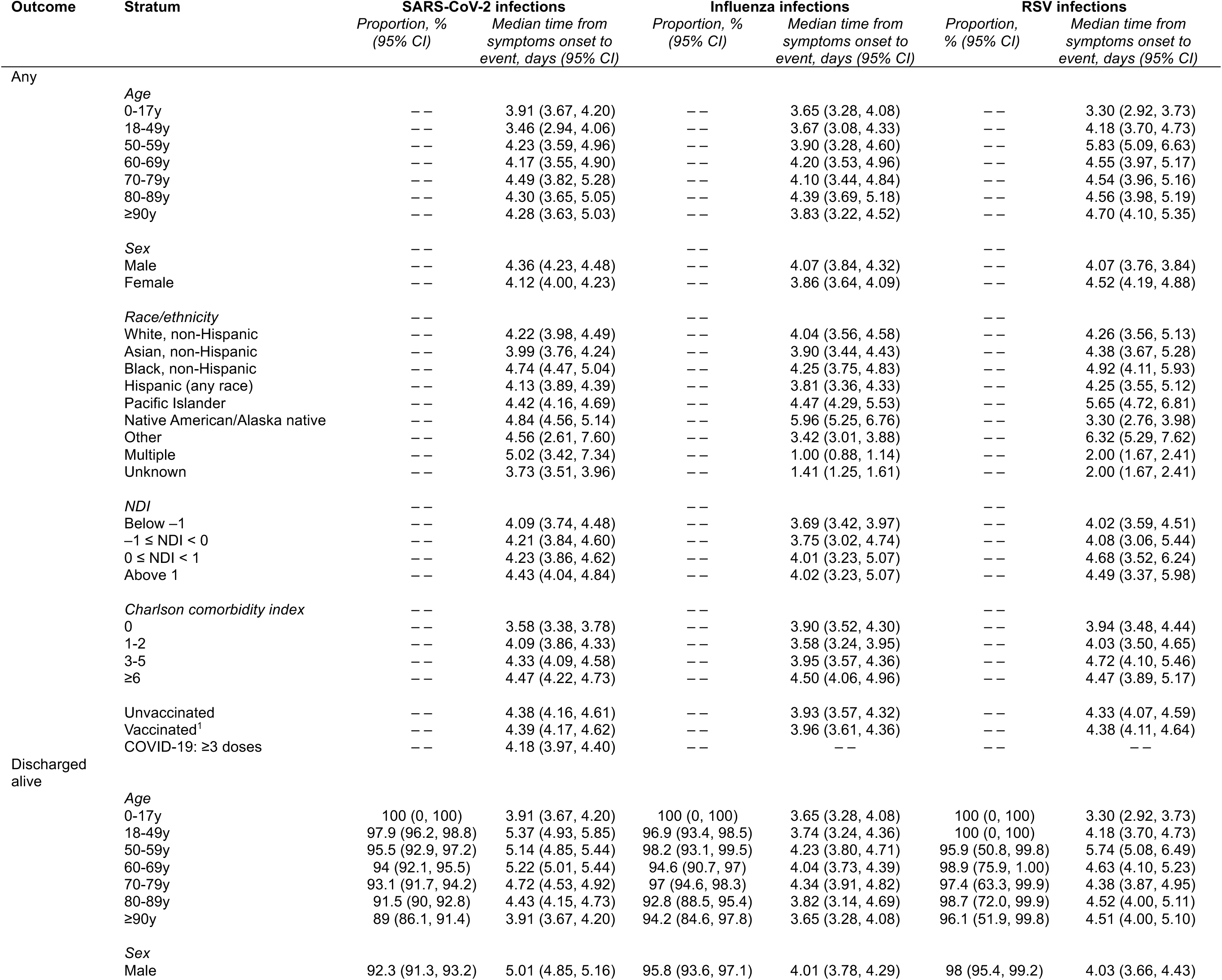
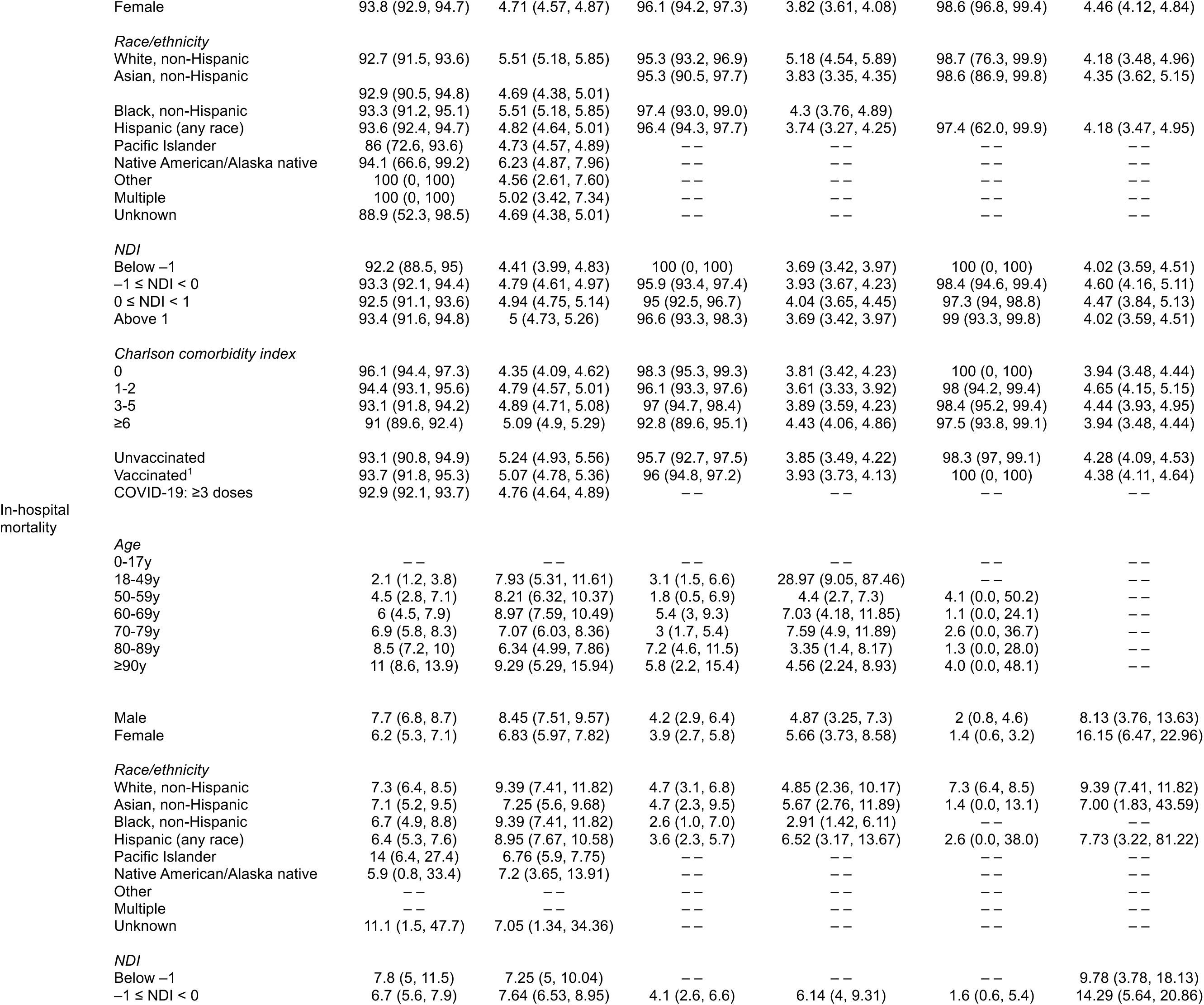
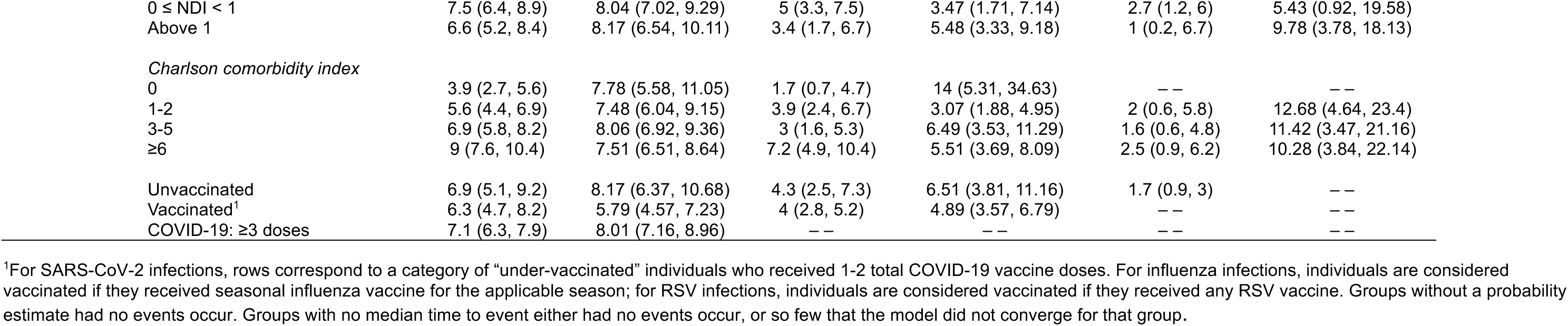
Hospital length of stay estimates for admissions associated with SARS-CoV-2 infection leading to discharge or mortality.

**Figure S1:**
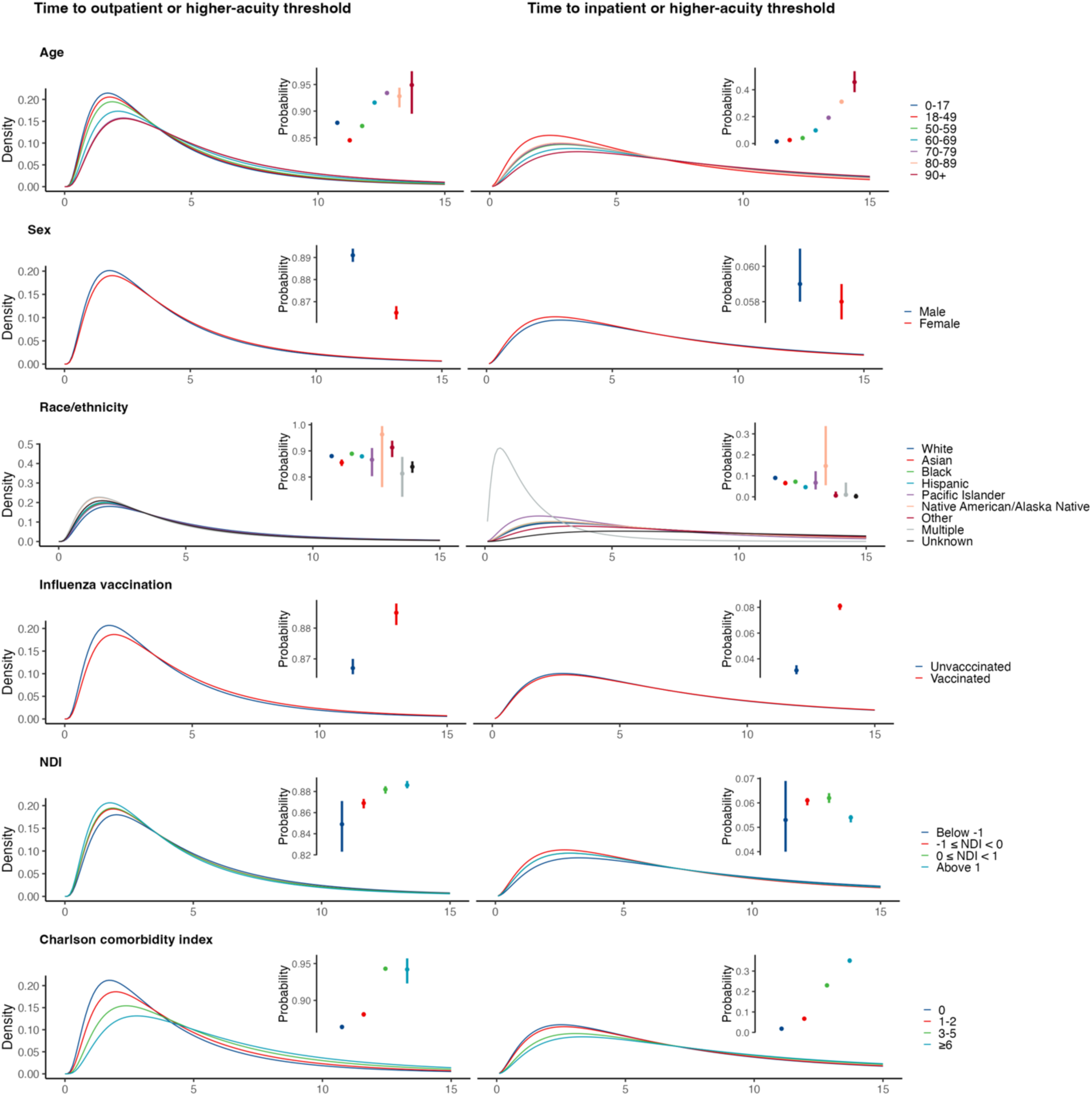
Probabilities and time-to-event distributions of reaching outpatient and inpatient acuity thresholds from symptom onset for illness associated with influenza infection across demographic subgroups. We illustrate best-fitting distributions for times to the indicated events from symptom onset. Probabilities of reaching acuity thresholds are available in **Tables S8**-**S12**.

**Figure S2:**
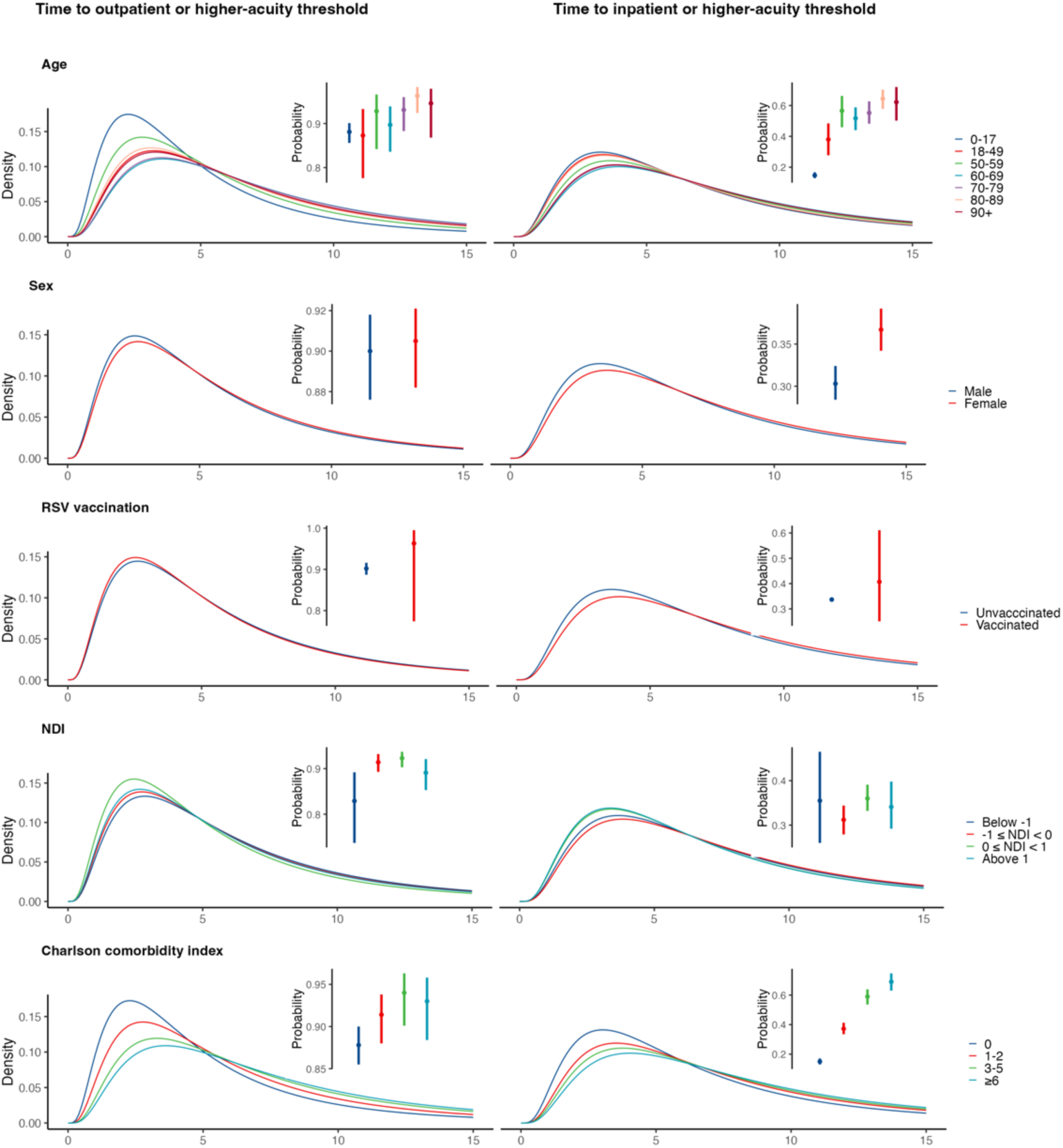
Probabilities and time-to-event distributions of reaching outpatient and inpatient acuity thresholds from symptom onset for illness associated with RSV infection across demographic subgroups. We illustrate best-fitting distributions for times to the indicated events from symptom onset. Probabilities of reaching acuity thresholds are available in **Tables S8-S12**

## REFERENCES

1. Hanage WP, Schaffner W (2025) Burden of acute respiratory infections caused by influenza virus, respiratory syncytial virus, and SARS-CoV-2 with consideration of older adults: A narrative review. Infectious Diseases and Therapy 14(1):5–37.

2. Putri WCWS, Muscatello DJ, Stockwell MS, Newall AT (2018) Economic burden of seasonal influenza in the United States. Vaccine 36(27):3960–3966.

3. Carrico J, Hicks KA, Wilson E, Panozzo CA, Ghaswalla P (2024) The annual economic burden of respiratory syncytial virus in adults in the United States. Journal of Infectious Diseases 230(2):e342–e352.

4. Loo SL, Howerton E, Contamin L, Smith CP, Borchering RK, Mullany LC, et al. (2024) The US COVID-19 and Influenza Scenario Modeling Hubs: Delivering long-term projections to guide policy. Epidemics 46:100738.

5. Angulo FJ, Finelli L, Swerdlow DL (2021) Estimation of US SARS-CoV-2 infections, symptomatic infections, hospitalizations, and deaths using seroprevalence surveys. JAMA Network Open 4(1):e2033706.

6. Salje H, Tran Kiem C, Lefrancq N, Courtejoie N, Bosetti P, Paireau J, et al. (2020) Estimating the burden of SARS-CoV-2 in France. Science 369(6500):208–211.

7. Lewnard JA, Liu VX, Jackson ML, Schmidt MA, Jewell BL, Flores JP, et al. (2020) Incidence, clinical outcomes, and transmission dynamics of severe coronavirus disease 2019 in California and Washington: Prospective cohort study. BMJ 369:m1923.

8. Rees EM, Nightingale ES, Jafari Y, Waterlow NR, Clifford S, Pearson CAB, et al. (2020) COVID-19 length of hospital stay: A systematic review and data synthesis. BMC Medicine 18(1):270.

9. Rolfes MA, Foppa IM, Garg S, Flannery B, Brammer L, Singleton JA, et al. (2018) Annual estimates of the burden of seasonal influenza in the United States: A tool for strengthening influenza surveillance and preparedness. Influenza and Other Respiratory Viruses 12(1):132–137.

10. Presanis AM, De Angelis D, Team TNYCSFI, Hagy A, Reed C, Riley S, et al. (2009) The severity of pandemic H1N1 influenza in the United States, from April to July 2009: A Bayesian analysis. PLOS Medicine 6(12):e1000207.

11. Reed C, Angulo FJ, Swerdlow DL, Lipsitch M, Meltzer MI, Jernigan D, et al. (2009) Estimates of the prevalence of pandemic (H1N1) 2009, United States, April–July 2009. Emerging Infectious Diseases 15(12):2004–2007.

12. Koebnick C, Langer-Gould AM, Gould MK, Chao CR, Iyer RL, Smith N, et al. (2012) Sociodemographic characteristics of members of a large, integrated health care system: Comparison with US Census Bureau data. The Permanente Journal 16(3):37–41.

13. Davis AC, Voelkel JL, Remmers CL, Adams JL, McGlynn EA (2023) Comparing Kaiser Permanente members to the general population: Implications for generalizability of research. The Permanente Journal 27(2):87–98.

14. Malden DE, Tartof SY, Ackerson BK, Hong V, Skarbinski J, Yau V, et al. (2022) Natural language processing for improved COVID-19 characterization: Evidence from more than 350,000 patients in a large integrated health care system. JMIR Public Health Surveillance 8:e41529.

15. Jackson C (2016) flexsurv: A platform for parametric survival modeling in R. Journal of Statistical Software 70:1– 33.

16. Messer LC, Laraia BA, Kaufman JS, Eyster J, Holzman C, Culhane J, et al. (2006) The development of a standardized neighborhood deprivation index. Journal of Urban Health 83(6):1041–1052.

17. Alahmadi A, Belet S, Black A, Cromer D, Flegg JA, House T, et al. (2020) Influencing public health policy with data-informed mathematical models of infectious diseases: Recent developments and new challenges. Epidemics

18. De Angelis D, Presanis AM, Birrell PJ, Tomba GS, House T (2015) Four key challenges in infectious disease modelling using data from multiple sources. Epidemics 10:83–87.

19. Lewnard JA, Reingold AL (2019) Emerging challenges and opportunities in infectious disease epidemiology. American Journal of Epidemiology 188(5):873–882.

20. Howerton E, Contamin L, Mullany LC, Qin M, Reich NG, Bents S, et al. (2023) Evaluation of the US COVID-19 Scenario Modeling Hub for informing pandemic response under uncertainty. Nature Communications 14(1):7260.

21. Mathis SM, Webber AE, León TM, Murray EL, Sun M, White LA, et al. (2024) Evaluation of FluSight influenza forecasting in the 2021–22 and 2022–23 seasons with a new target laboratory-confirmed influenza hospitalizations. Nature Communications 15(1):6289.

22. Wang D, Hu B, Hu C, Zhu F, Liu X, Zhang J, et al. (2020) Clinical characteristics of 138 hospitalized patients with 2019 novel coronavirus–infected pneumonia in Wuhan, China. JAMA 323(11):1061–1069.

23. Zhang J, Litvinova M, Wang W, Wang Y, Deng X, Chen X, et al. (2020) Evolving epidemiology and transmission dynamics of coronavirus disease 2019 outside Hubei province, China: A descriptive and modelling study. The Lancet Infectious Diseases 20(7):793–802.

24. Faes C, Abrams S, Van Beckhoven D, Meyfroidt G, Vlieghe E, Hens N, et al. (2020) Time between symptom onset, hospitalisation and recovery or death: Statistical analysis of Belgian COVID-19 patients. International Journal of Environmental Research and Public Health 17(20):7560.

25. Jackson CH, Grosso F, Kunzmann K, Corbella A, Gramegna M, Tirani M, et al. (2022) Trends in outcomes following COVID-19 symptom onset in Milan: A cohort study. BMJ Open 12(3):e054859.

26. Lewnard JA, Hong VX, Patel MM, Kahn R, Lipsitch M, Tartof SY (2022) Clinical outcomes associated with SARS-CoV-2 Omicron (B.1.1.529) variant and BA.1/BA.1.1 or BA.2 subvariant infection in southern California. Nature Medicine 28:1933–1943.

27. Garibaldi BT, Fiksel J, Muschelli J, Robinson ML, Rouhizadeh M, Perin J, et al. (2021) Patient trajectories among persons hospitalized for COVID-19. Annals of Internal Medicine 174(1):33–41.

28. Surie D, Yuengling KA, DeCuir J, Zhu Y, Lauring AS, Gaglani M, et al. (2024) Severity of respiratory syncytial virus vs COVID-19 and influenza among hospitalized US adults. JAMA Network Open 7(4):e244954.

29. Arntzen VH, Fiocco M, Geskus RB (2024) Two biases in incubation time estimation related to exposure. BMC Infectious Diseases 24(1):555.

30. Park SW, Sun K, Champredon D, Li M, Bolker BM, Earn DJD, et al. (2021) Forward-looking serial intervals correctly link epidemic growth to reproduction numbers. Proceedings of the National Academy of Sciences 118(2):e2011548118.

31. Miller AC, Hannah LA, Futoma J, Foti NJ, Fox EB, D’Amour A, et al. (2022) Statistical deconvolution for inference of infection time series. Epidemiology 33(4):470.

32. Ko JY, Danielson ML, Town M, Derado G, Greenlund KJ, Kirley PD, et al. (2021) Risk factors for coronavirus disease 2019 (COVID-19)–associated hospitalization: COVID-19–Associated Hospitalization Surveillance Network and Behavioral Risk Factor Surveillance System. Clinical Infectious Diseases 72(11):e695–e703.

33. Tartof SY, Aliabadi N, Goodwin G, Slezak J, Hong V, Ackerson B, et al. (2024) Estimated vaccine effectiveness for respiratory syncytial virus–related lower respiratory tract disease. JAMA Network Open 7(12):e2450832.

34. Tartof SY, Slezak JM, Frankland TB, Puzniak L, Hong V, Ackerson BK, et al. (2024) Estimated effectiveness of the BNT162b2 XBB vaccine against COVID-19. JAMA Internal Medicine 184(8):932–940.

35. Lewnard JA, Bruxvoort KJ, Hong VX, Grant LR, Jódar L, Cané A, et al. (2022) Effectiveness of pneumococcal conjugate vaccination against virus-associated lower respiratory tract infection among adults: A case-control study. Journal of Infectious Diseases 227(4):498–511.

36. Crawford FW, Weiss RE, Suchard MA (2015) Sex, lies and self-reported counts: Bayesian mixture models for heaping in longitudinal count data via birth-death processes. Annals of Applied Statistics 9(2):572–596.

